# Ultra-Rapid Droplet Digital PCR Enables Intraoperative Tumor Quantification

**DOI:** 10.1101/2024.05.29.24308126

**Authors:** Zachary R. Murphy, Emilia C. Bianchini, Andrew Smith, Lisa I. Körner, Teresa Russell, David Reinecke, Yuxiu Wang, Matija Snuderl, Daniel A. Orringer, Gilad D. Evrony

## Abstract

The diagnosis and treatment of tumors often depends on molecular-genetic data. However, rapid and iterative access to molecular data is not currently feasible during surgery, complicating intraoperative diagnosis and precluding measurement of tumor cell burdens at surgical margins to guide resections. To address this gap, we developed Ultra-Rapid droplet digital PCR (UR-ddPCR), which can be completed in 15 minutes from tissue to result with an accuracy comparable to standard ddPCR. We demonstrate UR-ddPCR assays for the IDH1 R132H and BRAF V600E clonal mutations that are present in many low-grade gliomas and melanomas, respectively. We illustrate the clinical feasibility of UR-ddPCR by performing it intraoperatively for 13 glioma cases. We further combine UR-ddPCR measurements with UR-stimulated Raman histology intraoperatively to estimate tumor cell densities in addition to tumor cell percentages. We anticipate that UR-ddPCR, along with future refinements in assay instrumentation, will enable novel point-of-care diagnostics and the development of molecularly-guided surgeries that improve clinical outcomes.

## Introduction

Molecular-genetic information is critically important for managing cancer treatment, but it is not currently accessible in the operating room. The standard clinical workflow only provides this information several days or more than a week after surgery^1,2^. Intraoperative knowledge of the molecular-genetic subtype of a tumor would be useful in guiding the surgical approach, and it would also avoid delays in planning post-surgical treatment. For example, stratification of a brain tumor as a low-grade versus high-grade glioma may justify a different resection strategy^3^. Additionally, the surgical decision of where to stop resecting tissue on the tumor margins is critical, but there is currently no technology to quantify the tumor cell percentage or density in a sample on the timescale required for intraoperative surgical guidance. In most surgeries, the boundaries of a tumor are determined by frozen histology and/or gross visualization by the surgeon, but this is imperfect, because it does not rely on molecular information. Ultra-rapid, intraoperative genetic subtyping and measurement of tumor cell percentage and density would therefore fundamentally change the approach surgeons take in resecting tumors and enable them to establish more optimal surgical endpoints.

Many cancer types frequently harbor one or more clonal mutations (i.e., mutations present in all the cells of the tumor) that are characteristic of that cancer type. These clonal hotspot mutations could allow for the development of widely applicable, yet targeted intraoperative assays that both identify the molecular-genetic subtype of a tumor and quantify the tumor cell percentage of tissue samples to guide tumor resection. One such hotspot mutation in gliomas is the IDH1 R132H mutation that is both frequent, occurring in approximately 65% of low-grade gliomas, and one of the most important prognostic biomarkers for adult gliomas^4,5^. Another example is the BRAF V600E mutation that is frequently present in melanoma and also observed in many other tumor types, including thyroid, lung, ovarian, and some low-grade brain tumors^6,7^. Each of these hotspot mutations is usually clonal^8,9^, such that measuring the mutant DNA fraction of these mutations in a tumor sample quantifies the tumor cell percentage in that sample.

Notably, for gliomas and melanomas, as well as other tumor types, the completeness or extent of surgical resection is an important prognostic factor^10-12^. Unfortunately, judging the boundaries of many tumors during surgery can be extraordinarily challenging^13^. Despite the introduction of a host of diagnostic modalities developed to help surgeons establish optimal endpoints for surgery, none can provide a rapid, direct, and accurate assessment of tumor cell burden at surgical margins^12^. Additionally, for gliomas, the effects of surgery are more pronounced in IDH1-mutant than IDH1-wild type tumors^14^, so it would be greatly beneficial to tailor surgical objectives while taking IDH1 status into account. For these reasons, the IDH1 R132H and BRAF V600E mutations are prime candidates for an ultra-rapid intraoperative assay.

To be useful for clinical decisions in the operating room, any technology for molecularly subtyping a tumor and quantifying tumor cell percentage should be able to deliver results rapidly in < 20 minutes with high sensitivity and specificity. Notably, there is no current technology meeting these requirements. Ultra-rapid histological methods can achieve the requirements for efficient integration into the surgical workflow^15^, but their results rely primarily on indirect inference based on tumor cell morphology rather than on genetic features that provide definitive tumor subtyping and detection. Previously, Shankar, et al. proposed a targeted quantitative PCR assay, but the tissue-to-result time was ∼ 60 minutes^16^. More recently, Wadden et al. developed a targeted sequencing method for rapid measurement of tumor cell percentage in a single sample in ∼ 30 minutes^17^, which represents a significant advance, but this is still not sufficiently fast for repeated intraoperative use. Extreme PCR is a method for conducting PCR in < 30 seconds^18^, but to date, it has only been implemented as a bulk assay that is unable to sensitively determine mutant DNA fraction in a sample, which would be necessary for measuring tumor cell percentage. In contrast, droplet-digital PCR (ddPCR), in which a bulk PCR reaction is partitioned into thousands of nanoliter-sized reactions, can measure tumor cell percentage with high sensitivity and specificity, but it currently takes > 2 hours to perform, not including the time for extracting DNA from tissue.

Here, we present the first Ultra-Rapid (UR)-ddPCR method that achieves a total tissue-to-result time of 15 minutes (schematic in **Figure 1A**). Our method combines the speed of Extreme PCR and the sensitivity of standard ddPCR, along with a compatible ultra-rapid DNA extraction procedure. We developed UR-ddPCR assays for both the IDH1 R132H and BRAF V600E mutations. We further implement this technology to genetically subtype and measure tumor percentage during glioma surgeries—demonstrating the first ultra-rapid (< 20 minutes) genetic assay performed in an operating room.

**Figure 1.**
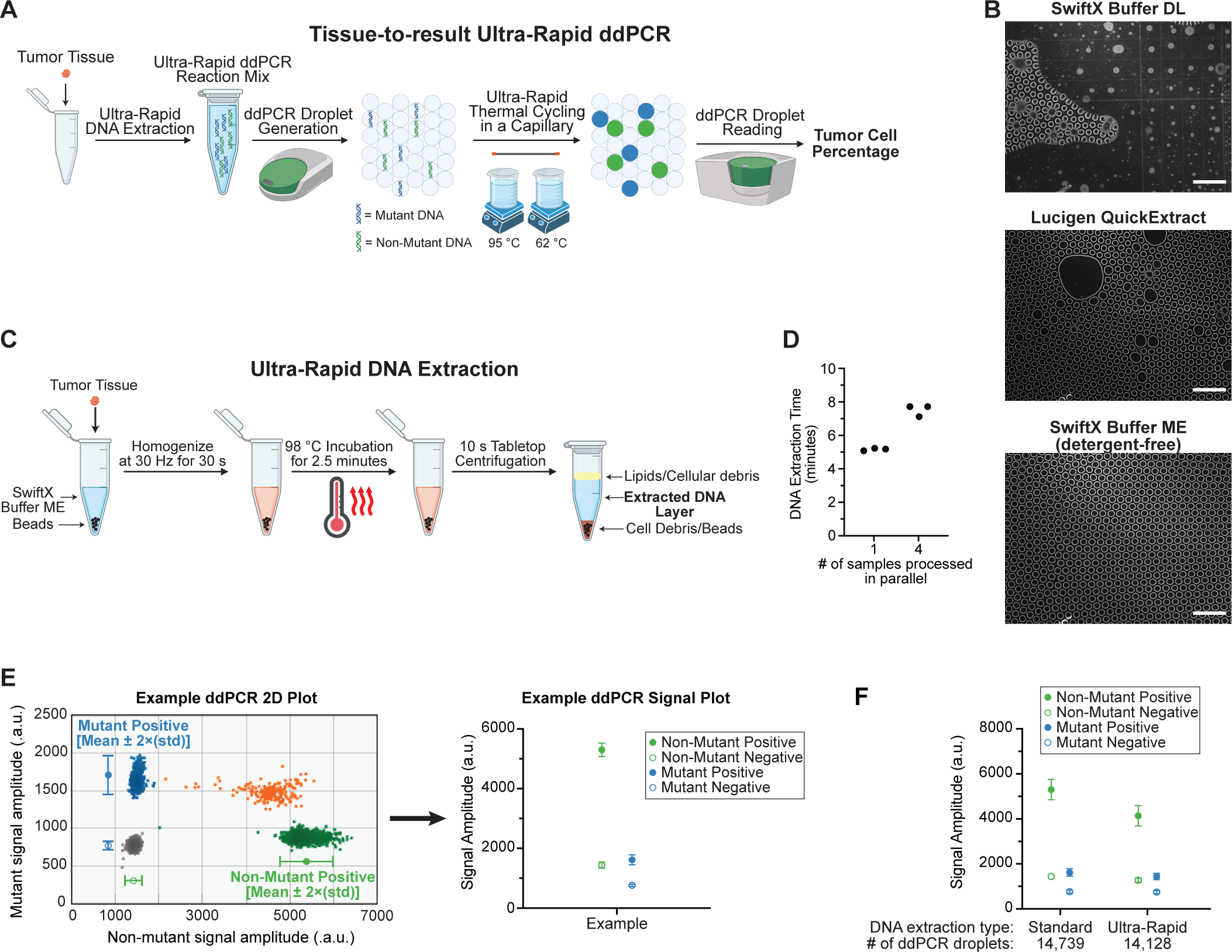
Process schematic and Ultra-Rapid DNA extraction. A) Schematic of Tissue-to-result Ultra-Rapid ddPCR. B) Microscopy images of ddPCR droplets after droplet generation from a reaction mix containing an additional 4 μL of each specified DNA extraction buffer. Scale bars, 100 µm. C) Schematic of Ultra-Rapid (UR) DNA extraction. s, seconds. D) The total time of UR-DNA extraction when processing 1 sample at a time (N=3 replicates) and 4 samples in parallel (N=3 replicates). E) Schematic of how ddPCR signal plots shown in the manuscript correspond to raw ddPCR 2-dimensional (2D) plots. ddPCR signal plots show the mean fluorescence signal amplitude (a.u., arbitrary units) of the double-negative population and each single-positive population with error bars of ± 2 standard deviations (std). Note that the signal level of the double-positive population (i.e., mutant^+^/non-mutant^+^) is not depicted in ddPCR signal plots. Additionally, we do not depict positive populations on ddPCR signal plots if they contain 3 or fewer droplets, as these cannot be distinguished from rare false positive events. F) ddPCR signal plot of a standard ddPCR assay for IDH1 R132H DNA with input from either a standard DNA extraction or UR-DNA extraction. The total number of ddPCR droplets measured for each assay are listed below each sample.

## Results

### Ultra-Rapid DNA Extraction

Standard DNA extraction from tissues typically requires more than 30 minutes to perform. Therefore, the first step to develop an Ultra-Rapid (UR) ddPCR assay feasible for intraoperative use was to create an UR-DNA extraction method that is compatible with ddPCR. This is challenging because nearly all DNA extraction lysis buffers contain detergents that interfere with ddPCR droplet formation and stability. We initially tested two commonly used rapid DNA extraction buffers—Buffer DL from the SwiftX DNA extraction kit and Lucigen QuickExtract solution—by adding them to a ddPCR reaction mix prior to droplet generation. However, we found that neither of these buffers was compatible with ddPCR droplet formation (**Figure 1B**). In contrast, a detergent-free DNA extraction buffer, SwiftX Buffer ME, maintained ddPCR droplet integrity (**Figure 1B**). Using SwiftX Buffer ME, we then proceeded to develop an UR-DNA extraction method with a goal of 5 minutes per sample.

For the first step of UR-DNA extraction, we utilized bead homogenization in the presence of SwiftX Buffer ME to rapidly lyse cells, and we found that bead homogenization for only 30 seconds was sufficient to fully homogenize control brain tissue. We then incubate the homogenate at 98 °C for 2.5 minutes, during which SwiftX Buffer ME liberates DNA from associated proteins. We observed that a subsequent brief 10-second centrifugation efficiently separates cellular debris from the clarified lysate containing DNA. Our final UR-DNA extraction protocol comprised of these three brief steps—bead homogenization, heat incubation, and centrifugation—achieved our goal of 5 minutes for processing one tissue sample (**Figures 1C,D** and **Methods**).

We confirmed the compatibility of our UR-DNA extraction protocol with ddPCR by performing a standard ddPCR assay with input from either a standard DNA extraction or an UR-DNA extraction. UR-DNA extraction slightly reduced the ddPCR signal level and the separation between positive and negative droplet populations, but it did not affect the total number of ddPCR droplets (**Figures 1E,F**). Additionally, our UR-DNA extraction method can be scaled to process four samples in parallel in 7 minutes (**Figure 1D**). Therefore, our UR-DNA extraction achieved the first necessary step in developing an intraoperative UR-ddPCR assay.

### Ultra-Rapid ddPCR Thermal Cycling

To enable intraoperative use, the total time of a tissue-to-result UR-ddPCR assay should be less than 20 minutes. However, standard ddPCR thermal cycling, the longest step of ddPCR, takes ∼ 2 hours. Therefore, UR-ddPCR required a drastic reduction in ddPCR thermal cycling time to less than 5 minutes, which combined with UR-DNA Extraction (∼ 5 minutes), droplet generation (∼ 3 minutes), and droplet reading (∼ 3 minutes) would total less than 20 minutes. Using IDH1 R132H as the target assay (**Supplementary Table 1**), we extensively optimized every aspect of ddPCR thermal cycling to achieve this goal.

We began reducing the ddPCR thermal cycling time by shortening or removing each step of the standard ddPCR protocol and observing the ddPCR signal level after each successive change. Removing the 10-minute enzyme inactivation step, reducing the denaturation time from 30 seconds to 1 second, and reducing the annealing/extension step from 60 seconds to 15 seconds either maintained or only slightly reduced the ddPCR signal level and the separation between droplet populations (**Figure 2A**). However, when we removed the 10-minute heat activation step, the ddPCR signal was completely lost (**Figure 2A**). To rescue the ddPCR signal in the absence of a heat activation step, we added an aptamer-inhibited hot-start Taq polymerase (Aptamer HS Taq) to the ddPCR reaction mix that is immediately activated above 45 °C, in contrast to the prolonged heat activation step required by the standard ddPCR polymerase. Aptamer HS Taq allowed us to eliminate the heat activation step while retaining the ability to hydrolyze the probes to generate signal (**Figure 2A**).

**Figure 2.**
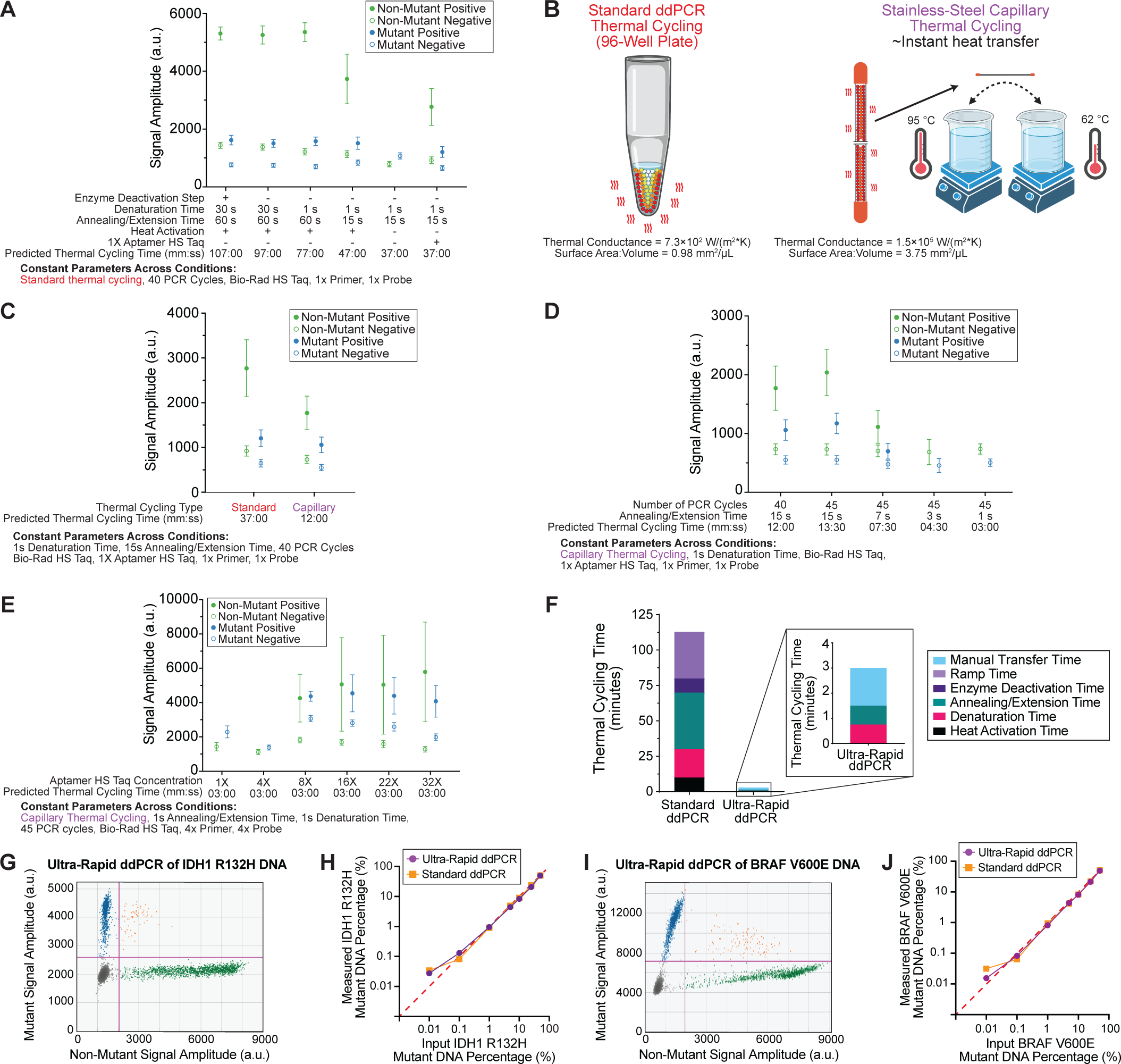
Ultra-Rapid ddPCR thermal cycling. A) ddPCR signal plot for the IDH1 R132H assay with multiple thermal cycling and ddPCR reaction mix conditions that reduce the total thermal cycling time (m, minutes; s, seconds). All modified parameters are listed for each sample, and the parameters that were constant for all samples are listed in the “Constant Parameters Across Conditions” section. 1x primer and 1x probe concentrations refer to the concentrations used in standard ddPCR, and the 1x Aptamer Hot-Start (HS) Taq concentration refers to 0.025 U/μL (**Methods**). The modified and constant parameters for all the other ddPCR signal plots in this figure are described in the same manner. a.u., arbitrary units. B) A schematic comparison of the properties of a standard ddPCR 96-well plate’s well and a stainless-steel capillary with a depiction of the capillary thermal cycling process. See the **Supplementary Note** for calculation details. C) ddPCR signal plot for the IDH1 R132H assay showing the effect of switching to the stainless-steel capillary and water bath thermal cycling system. D) ddPCR signal plot for the IDH1 R132H assay showing the effect of decreasing the annealing/extension time from 15 seconds to 1 second. E) ddPCR signal plot for the IDH1 R132H assay showing the effect of increasing the Aptamer HS Taq concentration, in the presence of higher 4x primer and probe concentrations. F) Schematic of the estimated standard ddPCR and UR-ddPCR thermal cycling times. The standard ddPCR bar depicts the times of each standard ddPCR step. The thermocycler ramp time was calculated as the difference between the total PCR run time and the sum of the times of individual thermal cycling steps. The UR-ddPCR bar depicts the time of the denaturation and annealing/extension steps (1s per cycle) and typical time required for manually transferring the capillary between water baths (1s per water bath transfer; 2 transfers per cycle). G) Representative two-dimensional ddPCR plot from an IDH1 R132H UR-ddPCR assay of DNA extracted from an IDH1 R132H mutant tumor with standard DNA extraction. H) Sensitivity of the IDH1 R132H assay by UR-ddPCR compared to standard ddPCR with DNA inputs containing known percentages of IDH1 R132H mutant DNA. The red line (y = x) depicts theoretical identical measurements by standard ddPCR and UR-ddPCR. This experiment used 32X Aptamer HS Taq concentration. See **Supplemental** Figure 1D for data of an experiment using 25X Aptamer HS Taq concentration. I) Representative two-dimensional ddPCR plot from a BRAF V600E UR-ddPCR assay of 50% BRAF V600E mutant reference DNA. J) Sensitivity of the BRAF V600E assay by UR-ddPCR compared to standard ddPCR with DNA inputs that containing known percentages of BRAF V600E mutant DNA. This experiment used 25X Aptamer HS Taq concentration. The red line depicts the theoretical exact measurement of the input mutant DNA percentage.

Collectively, the above changes reduced the total thermal cycling time from 107 minutes to 37 minutes, but 25 minutes of this remaining time was due to the time spent by the thermal cycler instrument ramping between temperatures. Since even thermal cycler instruments with the most rapid ramping speeds would not allow us to achieve our desired total tissue-to-result ddPCR time, we switched from a standard thermal cycler instrument utilizing polypropylene reaction plates to a stainless-steel capillary water bath thermal cycling system (**Figure 2B**)^18^. The stainless-steel capillary both increases the surface area to volume ratio of the reaction 3.8-fold and increases the thermal conductance approximately 204-fold relative to a standard ddPCR reaction (**Figure 2B** and **Supplementary Note**). This stainless-steel capillary system reduced the total ramping time to 1.5 minutes and was limited only by the speed of the technician moving the capillary between the water baths. Combined with the prior protocol changes, this reduced the total thermal cycling time to 12 minutes while maintaining significant ddPCR signal level and droplet population separation (**Figure 2C**).

The last challenge in decreasing the thermal cycling time was to further reduce the annealing/extension step to less than 15 seconds. We attempted to shorten the annealing/extension step without further modification of the ddPCR reaction mix, first by increasing from 40 to 45 PCR cycles, but we could not achieve ddPCR droplet population separation with annealing/extension times below 7 seconds (**Figure 2D**). Extreme PCR, a method that can complete a full PCR cycling program in less than 30 seconds, achieves this by significantly increasing the concentration of the primers and polymerase^18^. Applying this principle to our ddPCR reaction, we were able to reduce the annealing/extension step to only 1 second by increasing the number of PCR cycles to 45, increasing the Aptamer HS Taq concentration 32-fold, and increasing the concentrations of primers and probes 4-fold (**Figure 2E** and **Supplementary** Figure 1). This yielded an unprecedented total ddPCR thermal cycling time of 3 minutes (**Figure 2F**), which meets the time constraints necessary for an intraoperative tissue-to-result UR-ddPCR assay.

To confirm that UR-ddPCR maintained the high sensitivity and specificity of standard ddPCR, we assayed DNA standards with known IDH1 R132H mutant DNA percentages between 50% and 0.01% using both standard and UR-ddPCR. We found that both standard and UR-ddPCR assay accurately measured the mutant DNA percentage down to 0.1% and had false-positive rates of approximately 0.05% (**Figures 2G,H**). In this experiment, the percentage of droplets that were positive for either mutant or non-mutant DNA was lower in UR-ddPCR (15%) than in standard ddPCR (29%). This suggests that UR-ddPCR does not amplify all the droplets containing target DNA, but that mutant and non-mutant droplets amplify proportionally such that the mutant DNA percentage is accurate and concordant with standard ddPCR down to a level of 0.1% mutant DNA. Altogether, these results demonstrate that UR-ddPCR provides ultra-rapid speed without sacrificing the high sensitivity and specificity of standard ddPCR.

In addition to the IDH1 R132H mutation, we developed an UR-ddPCR assay for the BRAF V600E mutation (**Supplementary Table 1 and Supplementary** Figure 2). After optimization, this assay achieved similar signal separation as the IDH1 R132H UR-ddPCR assay with similar conditions except it uses fewer PCR cycles (40) and longer annealing/extension time (5 seconds) for a total thermal cycling time of 6 minutes (**Figure 2I**). The BRAF V600E UR-ddPCR assay also matched the sensitivity and specificity of standard ddPCR by measuring mutant DNA percentage down to 0.1% (**Figure 2J**), with a false-positive rate of approximately 0.04%. These results demonstrate that UR-ddPCR is generalizable to other hotspot mutations.

### Tissue-to-Result Ultra-Rapid ddPCR in the Laboratory

Next, we combined our UR-DNA extraction and UR-ddPCR in a streamlined tissue-to-result UR-ddPCR (**Methods**). We first tested this process in the laboratory on 15 tumor samples obtained from two IDH1 R132H-mutant oligodendrogliomas. We profiled these 15 samples in six experiments: three experiments profiling one sample at a time, and three experiments each profiling four samples in parallel.

In every experiment profiling these samples, tissue-to-result UR-ddPCR achieved ddPCR signal levels similar to our prior UR-ddPCR profiling of purified DNA (**Figures 2G and 3A**). Additionally, mutant DNA percentages measured by tissue-to-result UR-ddPCR and by standard ddPCR of the same UR-DNA extraction lysates were highly concordant (**Figure 3B**). Only 2 of the 15 samples showed a statistically significant difference between UR-ddPCR and standard ddPCR mutant DNA percentage measurements and the absolute differences in these measurements were only 2.7% and 3.2% (**Figure 3B** and **Supplementary Table 2**). These tissue-to-result UR-ddPCR results were achieved in an average of 15 minutes and 20 seconds when profiling one sample at a time (N=3 experiments) and an average of 27 minutes and 25 seconds when testing 4 samples in parallel (N=3 experiments) (**Figure 3C**). These results indicate that tissue-to-result UR-ddPCR can identify a tumor genetic subtype and quantify mutant DNA percentage with high accuracy and ultra-rapid speed.

**Figure 3.**
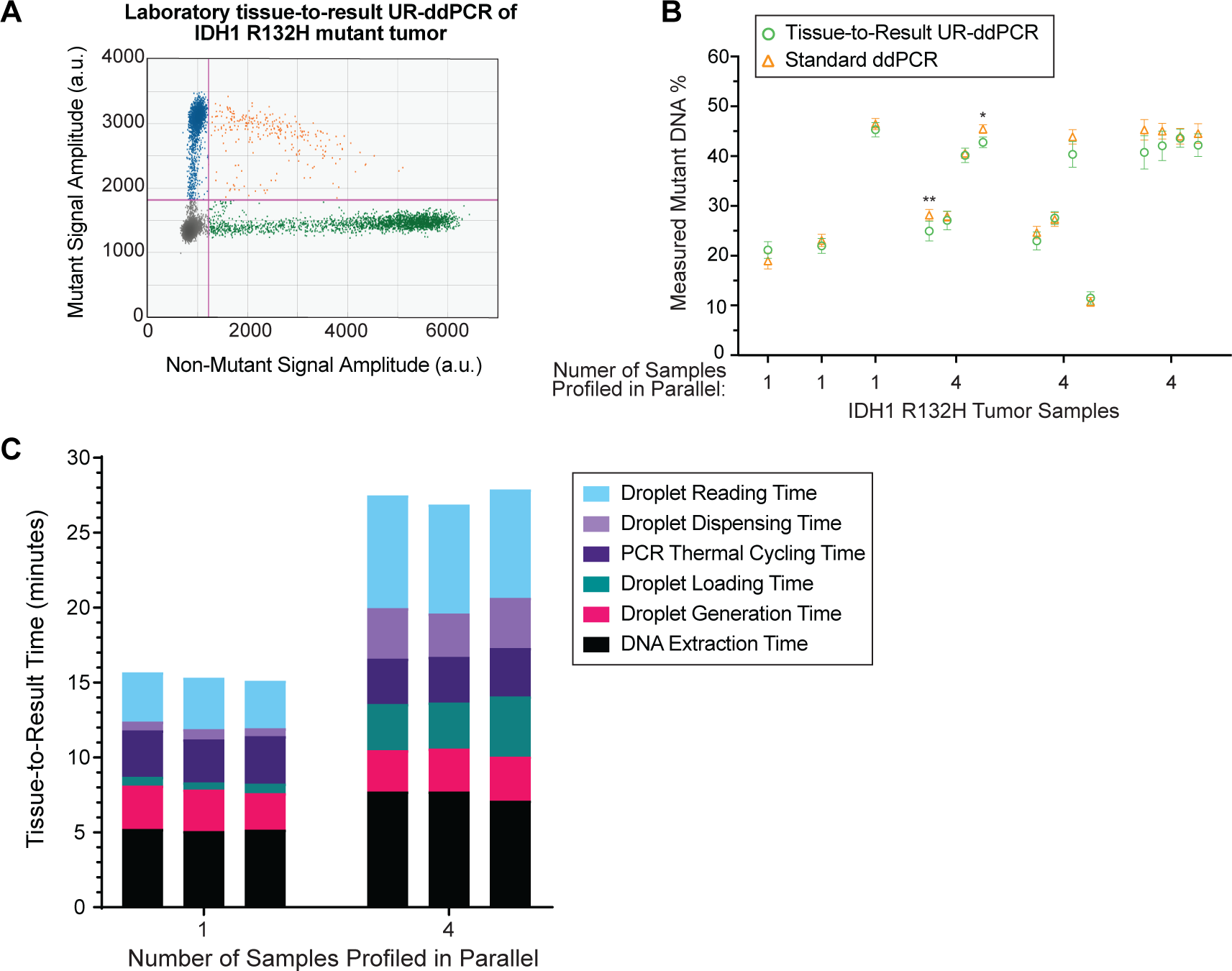
Tissue-to-result Ultra-Rapid ddPCR in the laboratory. A) Representative two-dimensional ddPCR plot of a tissue-to-result UR-ddPCR assay of an IDH1 R132H mutant tumor tissue sample. a.u., arbitrary units. B) Mutant DNA percentages of IDH1 R132H mutant tumor samples measured by tissue-to-result UR-ddPCR compared to standard ddPCR of the same UR-DNA extraction extracts. In each experiment, tumor samples were profiled either one at a time or four samples in parallel. Error bars, 95% Poisson confidence intervals. Significant differences: p < 0.05 (*), p< 0.005 (**), as measured by the Poisson difference of means test. C) The step-by-step time of the tissue-to-result UR-ddPCR assays shown in Figure 3B.

### Intraoperative Ultra-Rapid ddPCR

Since our ultimate goal in developing tissue-to-result UR-ddPCR was its use as a real-time guide for surgeons during operations, we implemented it in the operating room in 13 adult glioma cases. Note that we subsequently refer to tissue-to-result UR-ddPCR as only UR-ddPCR. We first designed an efficient layout on mobile carts of all the items required for UR-ddPCR (**Supplementary** Figure 3**, Supplementary Table 3, and Methods**). To fully assess UR-ddPCR performance, we profiled every sample with both UR-ddPCR and then with standard ddPCR after the operation was over. Additionally, on each day in the operating room, we confirmed our assay’s performance with two negative control assays, one without DNA input and one with non-mutant DNA input, and with a positive control assay of control DNA with a known mutant DNA percentage (**Methods**).

Each tissue sample provided by the surgeon for intraoperative diagnostics (average specimen size ∼ 8 x 8 x 2 mm) in the operating room was first split into approximately two halves: one half for UR-Stimulated Raman Histology^15^ (NIO system), which images the tissue in 5 minutes, followed by neuropathology analysis, and the other half for parallel profiling by UR-ddPCR (**Figure 4A**).

**Figure 4.**
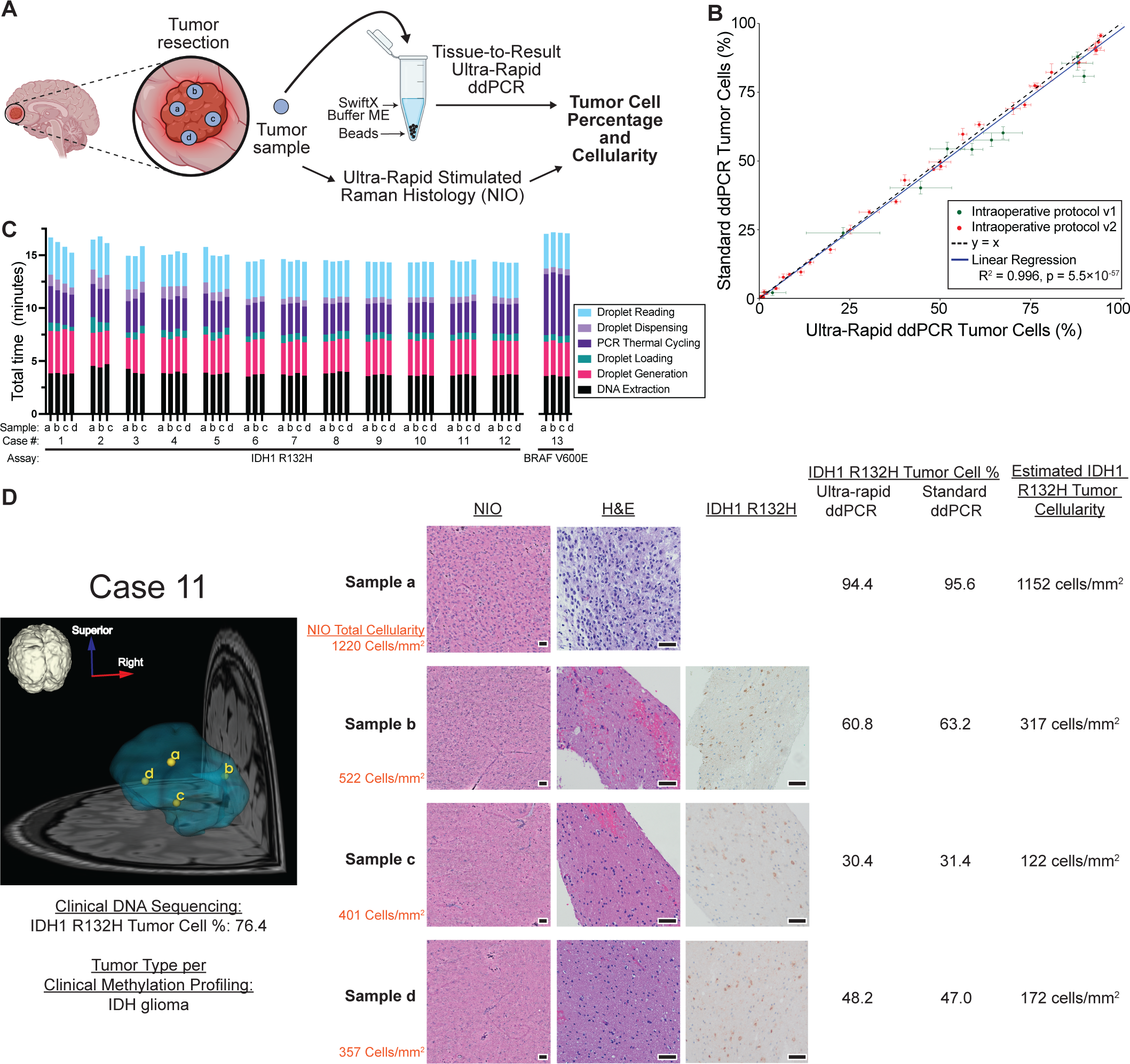
Intraoperative Ultra-Rapid ddPCR. A) Schematic of intraoperative tissue-to-result UR-ddPCR. B) Intraoperative tumor cell percentages measured by tissue-to-result UR-ddPCR compared to measurements of the same samples by standard ddPCR. We colored separately samples assayed with intraoperative UR-ddPCR protocols version 1 (v1) versus version 2 (v2), which differ in how the UR-ddPCR reaction mix is prepared (**Methods**). Error bars, 95% Poisson confidence intervals. See **Supplementary Table 4** for data of each sample and **Supplementary** Figures 4A,B for tumor cell percentage and estimated tumor cellularity plots of each sample grouped by surgical case. C) The step-by-step time of all intraoperative tissue-to-result UR-ddPCR cases. D) Comprehensive data of one representative operating room case (case 11), including anatomical tumor (green) and sample (yellow) locations overlaid on the subject’s brain MRI, clinical genome sequencing results, methylation profiling results, NIO UR-stimulated Raman histology that calculates total cellularity, H&E histology, IDH1 R132H staining (not available for samples ‘a’), tissue-to-result UR-ddPCR and standard ddPCR cell percentage measurements, and estimated IDH1 R132H tumor cellularity that is calculated as [NIO total cellularity] x [tissue-to-result UR-ddPCR tumor cell percentage]. Scale bars (black bar width), 50 µm. See **Supplementary Table 4** for details and **Supplementary File 1** for similar figures for all operating room cases.

We assayed multiple samples per case, including core and tumor margin samples, for a total of 49 samples across the 13 surgical cases (**Supplementary Table 4**). Tumor cell percentages measured by intraoperative UR-ddPCR were highly concordant with standard ddPCR performed on the same sample lysates (**Figure 4B**). These UR-ddPCR results were achieved in an average of 14 minutes and 54 seconds for IDH1 R132H assays (N=45 samples, each processed individually) and an average of 17 minutes and 6 seconds for the BRAF V600E assays (N=4 samples, each processed individually) (**Figure 4C**). These results demonstrate that intraoperative UR-ddPCR can both identify tumor genetic subtype and quantify tumor cell percentage with high accuracy and ultra-rapid speed.

Since simulated Raman histologic images acquired intraoperatively in minutes provided us real-time measurements of total cellularity (cells/mm^2^), we were also able to multiply these measurements with the UR-ddPCR measurements of tumor cell percentage to obtain ultra-rapid estimates of mutant tumor cellularity (i.e., tumor cells/mm^2^). Notably, individual cases had a wide range of tumor cell percentage and tumor cellularity estimates, consistent with the wide dynamic range of our assay (**Supplementary Table 4 and Figures 2H,J**). For example, in case 5, we measured tumor cell percentages of 50.0%, 76.2%, 76.8%, and 4.6% for a core tumor sample and 3 different margin samples, respectively, which combined with UR-histology cellularity measurements estimated 282, 722, 546, and 12 tumor cells/mm^2^, respectively. Subsequent clinical sequencing of all cases was concordant with our UR-ddPCR results, including one IDH non-mutant tumor and one IDH2 mutant tumor that tested negative in our IDH1 R132H assay (**Supplementary Table 4**). We illustrate UR-ddPCR results together with stereotactic biopsy coordinates, UR-histology, standard histology, immunohistochemistry, and clinical molecular testing of a representative case in **Figure 4D** and of all other cases in **Supplementary File 1**. Overall, our workflow demonstrates an unprecedented ability to map tumor cell content during surgery.

## Discussion

Surgeons synthesize anatomic, physiologic, radiographic, and histologic data to create an operative strategy during tumor resection. To date, however, there has been no intraoperative method to rapidly and iteratively utilize tumor-specific genetic alterations to guide surgical resections. Capitalizing on clonal hotspot mutations that define many human malignancies^19^, we envisioned a strategy to streamline intraoperative diagnosis and enhance the precision of tumor resection through ultra-rapid ddPCR. Here, we developed and validated the first ultra-rapid ddPCR technology that achieves these goals, with detection down to 0.1% tumor cell percentage and < 1 tumor cell per mm^2^ of tissue in 15 minutes for a single sample, and in under 30 minutes for 4 samples profiled in parallel. Importantly, we validated the use of this technology in the operating room, demonstrating the fastest-reported intraoperative quantification of tumor cells.

Existing molecular methods proposed for intraoperative use include methylation profiling^20^, quantitative PCR^16^, targeted DNA sequencing^17^, and Crispr-Cas12a assays^21^. While intraoperative methylation profiling can distinguish a wide range of tumor subtypes, it takes 40 minutes to perform, it cannot classify samples with low tumor purity, and the technology has limited range and accuracy in quantifying tumor cell percentage^20^. Quantitative PCR, which has been demonstrated intraoperatively in ∼ 60 minutes from tissue to result^16^, also has limited accuracy compared to ddPCR^22^. Targeted DNA sequencing can quantify tumor cell percentage in a single tumor sample in ∼ 30 minutes^17^, but this time to result is not rapid enough for repeated use in the same surgical case. Crispr-Cas12a assays take ∼ 60 minutes and provide only a qualitative measurement of tumor cell burden^21^. In contrast to these methods, our tissue-to-result UR-ddPCR is the first approach that provides both quantitative measurement of tumor cell burdens and a speed sufficient for repeated use in the operating room.

Our demonstration of UR-ddPCR assays for both IDH1 R132H and BRAF V600E hotspot mutations motivates extension of this technique to a broader array of clinically relevant genetic loci. Accordingly, we envision future comprehensive panels of hotspot mutations specific for a given cancer type that can be used in an ultra-rapid genetic subtyping screen at the beginning of a surgery to help determine the overall resection strategy. This would be facilitated by new ddPCR instruments that can multiplex up to 12 targets per reaction, though assays for other loci may require optimization or may not be feasible due to local sequence contexts. For example, a screen for the 4 most frequent IDH1 mutations (R132H, R132C, R132S, and R132G) would capture approximately 75% of all low-grade gliomas^23^, and other frequently mutated loci would encompass an even larger fraction of brain tumors^24^. If one of the hotspot mutations is detected in the initial genetic subtyping screen, that mutation could subsequently be profiled by UR-ddPCR to serially measure tumor cell percentages throughout the surgical resection. Nevertheless, while UR-ddPCR and other rapid genetic assays focus on hotspot mutations, future ultra-rapid whole-genome sequencing will need to be developed for tumors that do not harbor hotspot mutations.

UR-ddPCR is well-positioned to complement the growing array of surgical adjuncts developed to assess completeness of resection, including navigation, ultrasound, intraoperative magnetic resonance imaging, fluorescent markers^25-27^, and Raman- and AI-based methods^15,28^. Importantly, no existing modality can detect tumor cells with the granularity and speed of the UR-ddPCR workflow proposed here. With increasing interest in more complete surgical resections, methods that can definitively reveal tumor cell infiltration are critically important so that resection boundaries are safely maximized. Moreover, for brain tumors, as it becomes clear that the impact of surgical resection varies based on glioma molecular subtype^3^, there is a need for rapid and accurate molecular diagnostics at the time of surgery.

Even with the hypothesized benefits of rapid intraoperative molecular data, prospective trials comparing UR-ddPCR-driven surgical outcomes to existing methods for establishing the extent of tumor resection will be necessary to judge its value to patients. Although molecular-guided resection may improve outcomes for some tumor types, other tumor types may be diffusely infiltrative such that there is no therapeutic benefit. It will also be beneficial to conduct studies that evaluate whether an UR-ddPCR workflow can speed clinical management through expedited molecular diagnosis. Finally, quantification of tumor infiltration at the margins during the primary tumor resection may in the future inform stratification of patients to different targeted and chemotherapy treatments.

In its current form, UR-ddPCR requires further refinement to ensure its translation to the broader field of surgical oncology. Specifically, its current reliance on manual handling of samples would greatly benefit from automation. It is feasible that a single microfluidic chip may be able to perform the entire UR-ddPCR process after UR-DNA extraction—i.e., generating droplets, thermal cycling, and reading droplets—at greatly increased speed and sample parallelization. Microfluidic chips already exist that perform some or all of these steps, albeit none designed for ultra-rapid speed^29,30^. We estimate that an UR-ddPCR microfluidics chip could decrease the time required for profiling a single sample by more than half and may eliminate the time difference between profiling multiple samples in parallel versus a single sample.

The challenge of achieving rapid molecular diagnosis and optimal surgical margins extends well beyond the tumor types profiled in this study. We envision that our UR-ddPCR technology will spur the development of increasingly rapid and more comprehensive intraoperative assays that will transform the surgical resections of tumors. Notably, UR-ddPCR may also be extended in the future to other point-of care diagnostics such as infectious diseases. UR-ddPCR demonstrates the potential of emerging ultra-rapid molecular assays to create a new standard for point-of-care molecular diagnostics in medicine.

## Supplementary Information

<u>Supplementary Figure 1. Additional Ultra-Rapid ddPCR thermal cycling optimizations</u>. A) ddPCR signal plot of increasing numbers of PCR cycles. For this and subsequent figure panels, all modified parameters are listed under each sample and parameters that were constant for all samples are listed in the “Constant Parameters Across Conditions” section. a.u., arbitrary units; m, minutes; s, seconds. B) ddPCR signal plot of increasing primer concentration. C) ddPCR signal plot of increasing probe concentration. D) Sensitivity of the IDH1 R132H assay by UR-ddPCR compared to standard ddPCR with DNA inputs containing known percentages of IDH1 R132H mutant DNA. The red line (y = x) depicts theoretical identical measurements by standard ddPCR and UR-ddPCR. This experiment used 25X Aptamer HS Taq concentration. See Figure 2H for data of an experiment using 32X Aptamer HS Taq concentration.

<u>Supplementary Figure 2. Effect of probe chemistry on UR-ddPCR performance</u>. Comparison of UR-ddPCR fluorescence signal amplitudes when using PrimeTime locked nucleic acid versus Affinity Plus probe chemistries with a HEX fluorophore, for a BRAF (non-mutant sequence) probe assaying control DNA with the same protocol for each probe type. X-axis, droplets in order of instrument reading. a.u., arbitrary units.

<u>Supplementary Figure 3. Layouts of intraoperative tissue-to-result Ultra-Rapid ddPCR mobile carts</u>. A) Tissue processing table. B) Pre-PCR and droplet reader table. C) Post-PCR table.

Supplementary Figure 4. Intraoperative UR-ddPCR measurements of tumor cell percentages and cellularities, grouped by surgical case. (A) Tumor cell percentages measured by intraoperative tissue-to-result UR-ddPCR compared to standard ddPCR of the same samples. Error bars, 95% Poisson confidence intervals. (B) Estimated tumor cellularities calculated as [NIO total cellularity] x [tissue-to-result UR-ddPCR tumor cell percentage]. Cases are classified as mutant or non-mutant based on clinical sequencing. Error bars, 95% Poisson confidence intervals.

<u>Supplementary Table 1</u>. Primer and probe sequences.

<u>Supplementary Table 2</u>. IDH1 R132H DNA percentages measured in the laboratory by tissue-to-result UR-ddPCR and standard ddPCR of the same UR-DNA extraction lysates.

<u>Supplementary Table 3</u>. List of items required for intraoperative tissue-to-result UR-ddPCR.

<u>Supplementary Table 4</u>. Summary of intraoperative cases, including anatomical locations, clinical data, NIO (stimulated Raman histology) data, and UR- and standard ddPCR data. Note: clinical methylation and clinical genome sequencing data was profiled on different pieces of the tumor than those assayed by ddPCR, unless otherwise specified.

<u>Supplementary File 1</u>. MRI, clinical testing, histology, and ddPCR data for all intraoperative tissue-to-result UR-ddPCR cases and samples, except for the case illustrated in Figure 4D. Tumor and sample locations are colored in green and yellow, respectively, in MRI images. Note, some samples profiled by UR-ddPCR did not have corresponding histology data, and for some cases IDH1 R132H staining is shown below the MRI image because it was performed on a different clinical sample than the samples profiled by UR-ddPCR. The tissue-to-result UR-ddPCR tumor cell estimates are shown with the standard ddPCR tumor cell percentage from the same DNA extract. Estimated tumor cellularity was calculated as [NIO total cellularity] x [tissue-to-result UR-ddPCR tumor cell percentage]. Scale bars (black bar width), 50 µm. See **Supplementary Table 4** for further details.

<u>Supplementary Note</u>. Comparison of standard and ultra-rapid ddPCR thermal conductance and surface area to volume ratios.

## Methods

### Tissues for laboratory experiments

For laboratory experiments, we used tissues from three surgical resections of IDH1 R132H oligodendrogliomas of subjects consented under a human subjects protocol approved by the New York University Grossman School of Medicine Institutional Review Board. These three tumors were confirmed by clinical sequencing to have the IDH1 R132H mutation. One tumor sample was frozen fresh and initially stored at -80 °C, followed by standard DNA extraction (see below protocol) for use as positive control DNA for the IDH1 R132H standard ddPCR and UR-ddPCR laboratory experiments. The two other tumor samples were cut into small pieces (∼ 3 x 3 x 3 mm), each placed into separate 2 mL DNA LoBind tubes (Eppendorf), frozen fresh, and stored at -80 °C for later use in the tissue-to-result UR-ddPCR laboratory experiments.

Healthy cerebral cortex tissue (i.e., without the IDH1 R132H mutation) was obtained from the NIH NeuroBioBank (subject ID 5606, Broadman’s area 20), thawed, cut into small pieces (∼ 3 x 3 x 3 mm), placed into 2 mL DNA LoBind tubes, and frozen again at -80 °C for use as a negative control for the laboratory tissue-to-result UR-ddPCR experiments.

### Standard DNA extraction from tissues

Standard DNA extractions were conducted using the QIAamp DNA Mini Kit (Qiagen) per the manufacturer’s “DNA Purification from Tissues” protocol, including the optional RNase digestion step. DNA was eluted in 100 μL of 10 mM Tris pH 8 and stored at -20 °C. DNA quality was assessed with the NanoDrop One instrument (ThermoFisher) and quantified with the Qubit 1X dsDNA High Sensitivity Assay Kit (ThermoFisher).

### ddPCR assay design

IDH1 R132H and BRAF V600E ddPCR assay primers were designed with a combination of Primer3^31^ and manual design, and probes (5’ HEX or FAM fluorophores with 3’ Iowa Black quenchers) were designed by Integrated DNA Technologies (IDT). Primer and probe sequences are listed in **Supplementary Table 1**. The primers were designed to have short amplicons (< 150 base pairs) to reduce the time necessary for the polymerase to replicate the amplicon—thereby lowering the required annealing/extension time in PCR. Importantly, we found that in UR-ddPCR, PrimeTime locked nucleic acid probes (IDT) perform better than Affinity Plus probes (IDT) (**Supplementary** Figure 2), so we utilized the former in our experiments. Probes were designed with locked nucleic acids to enable shorter probes, which in turn increases the difference in melting temperatures for a matched versus mismatched base at the mutation location.

### Testing rapid DNA extraction buffer compatibility with ddPCR droplets

We tested the compatibility of rapid DNA extraction buffers, which may contain droplet-damaging detergents, with ddPCR droplets before developing the ultra-rapid DNA extraction. We tested the following three buffers: Buffer DL from the SwiftX DNA extraction kit (Xpedite Diagnostics), QuickExtract DNA Extraction Solution (Lucigen), and Buffer ME from the Swift X Media kit (Xpedite Diagnostics).

We added 4 μL of each rapid DNA extraction buffer to a standard ddPCR mix with a final volume of 22 μL containing: a) 1X ddPCR Supermix for Probes, no dUTP (Bio-Rad), b) 0.9 μM of each forward and reverse primer (IDT), c) 0.25 μM of each non-mutant and mutant probe (IDT), d) 0.23 U/μL of HaeIII (NEB). Standard droplet generation was conducted as described in the Standard ddPCR in the laboratory methods section, but instead of adding the droplets to a plate, 10 μL of the extract was added to a hemocytometer, covered with a coverslip, and imaged with light microscopy to determine the buffer’s compatibility with ddPCR droplets.

### Ultra-rapid DNA extraction in the laboratory

Ultra-rapid (UR) DNA extraction—the first step of tissue-to-result UR-ddPCR—was conducted for up to four samples in parallel.

To maximize speed, prior to the protocol we prepared the following: 1. We activated Buffer ME with Component P (Xpedite Diagnostics) by adding one 1 mL of Buffer ME to the tube containing dry Component P, pipetting thoroughly, transferring the full volume of resuspended Component P back into the Buffer ME bottle, mixing by inversion, and then aliquoting and storing the reagent at –20 °C; 2. We set the TissueLyser II (QIAGEN) instrument to a 30 Hz, 30 second program; 3. We pre-heated a ThermoMixer C (Eppendorf) to 98 °C; and, 4. For each sample, we pre-measured 0.1 grams of 0.2 mm RNase-free Stainless-Steel Beads (Next Advance) into 0.2 mL PCR tubes.

After completing these preparation steps, we added 200 μL of activated Buffer ME and the 0.1 grams of pre-measured 0.2 mm RNase-free Stainless-Steel Beads to the 2 mL DNA LoBind Tube containing the tumor tissue. We homogenized each sample with the TissueLyser II at 30 Hz for 30 seconds. Then, we incubated the post-homogenization mixture at 98 °C for 2.5 minutes on a ThermoMixer C to extract the DNA. Finally, this post-incubation lysate was briefly centrifuged for 10 seconds in an LSE Mini Microcentrifuge (Corning) to help separate cellular debris form layers containing the DNA used in ddPCR.

The resulting UR-DNA lysate usually separates into three layers: an opaque top layer, a clear middle layer, and a bottom layer containing the beads and cellular debris. DNA is present in the two top layers, and when the clear middle layer has sufficient volume and is clearly distinct from the top opaque layer, we use the middle layer for UR-ddPCR to minimize possible damage to droplets that can occur from cellular lipids in the opaque top layer. When the clear middle layer has insufficient volume or is not clearly distinct from the top opaque layer, we use the top opaque layer for UR-ddPCR. While the clear middle layer is preferable, DNA from either the clear or top opaque layers is compatible with UR-ddPCR.

### Standard ddPCR in the laboratory

The following protocol was used for standard ddPCR for both IDH1 R132H and BRAF V600E mutations:

For each sample, we added either 30 ng of genomic DNA or up to 0.5 μL of UR-DNA extract to a ddPCR mix with a final volume of 22 μL containing: a) 1X ddPCR Supermix for Probes, no dUTP (Bio-Rad), b) 0.9 μM of each forward and reverse primer (IDT), c) 0.25 μM of each non-mutant and mutant probe (IDT), and, d) 0.23 U/μL of HaeIII (NEB). 20 μL of this ddPCR mix was added to a DG8 droplet generation cartridge (Bio-Rad) sample well. We then added 20 μL of 1X ddPCR Buffer Control for Probes (Bio-Rad) to all unused DG8 cartridge sample wells and 70 μL of Droplet Generation Oil for Probes (Bio-Rad) to all DG8 cartridge oil wells. The DG8 cartridge was covered in a rubber gasket (Bio-Rad) and loaded in a QX200 Droplet Generator (Bio-Rad) to generate droplets. After generating droplets, we transferred 40 μL of the droplets from each sample to a ddPCR 96-well Plate (Bio-Rad, Cat. 12001925) and sealed the plate with a Pierceable Foil Heat Seal (Bio-Rad) and a PX1 PCR Plate Sealer (Bio-Rad).

The sealed ddPCR droplets were then thermal cycled on an Eppendorf Mastercycler X50L with the following protocol: 95 °C for 10 minutes, 45 cycles (IDH1 R132H) or 40 cycles (BRAF V600E) of 95 °C for 30 seconds and 62 °C for 1 minute, and 98 °C for 10 minutes. We read the amplified ddPCR droplets on a QX200 Droplet Reader (Bio-Rad) and analyzed the results with the QX Manager 1.1 Software. Thresholds for positive droplets were placed between droplet populations, and when separation between populations was not as clear, most droplets between populations (“rain” droplets) were included in the positive droplet population.

### Ultra-rapid ddPCR in the laboratory

Ultra-rapid (UR) ddPCR was developed via experiments that optimized the reagent concentrations and thermal cycling method, as detailed in the main text and figures. The final UR-ddPCR protocol, which is compatible with UR-DNA extraction, is described here.

Prior to performing UR-ddPCR, we prepared the following:

1. We prepared two 1-liter water baths, one at 95 °C and one at 62 °C. The water baths were heated on hot plates with continuously active magnetic stir bars, and temperatures were continuously measured with standard thermometers.
2. For each sample, we prepared a 20 μL (or 21.5 μL, if UR-DNA extract is assayed) reaction mix containing: a) 1X ddPCR Supermix for Probes, no dUTP, b) 3.6 μM of each forward and reverse primer, c) 1 μM of each non-mutant and mutant probe, d) 0.23 U/μL of HaeIII, and e) 0.8 U/μL (all laboratory UR-ddPCR experiments except for one of the IDH1 R132H ddPCR sensitivity experiments and the BRAF V600E sensitivity experiment) or 0.625 U/μL of Hot Start (aptamer-based) Taq DNA Polymerase (taken from a 20 U/μL stock that was made by diluting a 100 U/μL high-concentration preparation of polymerase custom ordered, catalog # M0495B-HC3, from NEB with 1x standard Taq buffer from NEB). As described in the text, the above primer and probe concentrations are 4x the concentrations used in standard ddPCR, and the Aptamer HS Taq polymerase is 32x (0.8 U/μL) or 25X (0.625 U/μL) relative to the 0.025 U/μL concentration recommended by NEB for standard PCR.
3. We added 20 μL of 1X ddPCR Buffer Control for Probes to all unused DG8 cartridge wells and 70 μL of Droplet Generation Oil for Probes (Bio-Rad) to all DG8 cartridge oil wells.
4. We initialized the QX Manager Software with the plate name, probes, and selected wells for analysis.

After completing these preparation steps, we added 2 μL of 15 ng/μL genomic DNA or 0.5 μL of UR-DNA extract to the reaction mix. ddPCR droplets were then generated as described above for standard ddPCR. However, after generating droplets, instead of loading the droplets into a 96-well plate, the droplets were loaded into thin-walled stainless steel capillaries obtained from either Ziggy’s Tubes and Wires (most experiments; product 18X304-CUT) or from Component Supply Company (product HTX-18X) with the following properties: 304 stainless steel, 18 gauge extra thin wall hypodermic tubing x 2.25 inches long, 0.0495 to 0.0505 inches outer diameter, 0.041 to 0.043 inches inner diameter, welded and drawn, burr-free. Note: after we discovered that some batches of capillaries cause droplets to be destroyed, we found that capillaries ordered from Ziggy’s Tubes and Wires, with instructions to undergo a standard wash followed by an ethanol wash, performed reliably. The droplets were loaded into the capillaries by loading a 200 μL pipette set to 45 μL with a pipette tip (Corning 4138). In this process, the pipette was loaded with a tip, then the capillary was manually inserted into the open end of the loaded pipette tip. The opposite end of the capillary not attached to the pipette tip was then put into the droplet generation cartridge at approximately a 45 °C angle and used to aspirate the droplets. The end of the capillary not attached to the pipette tip was then capped using a temperature-resistant silicone cap (92805K3, McMaster-Carr) that was pre-cut to 4 mm in length. Specifically, the cap was placed on the capillary by holding the pipette in one hand, then placing the capillary, while still attached to the pipette tip, between the middle and ring fingers of the other hand for support and carefully sliding the cap onto the tip of the capillary using the thumb and index fingers. The capillary was then gently detached from the pipette tip by holding the pipette vertically so that the capped end of the capillary was pointed downwards and pulling lightly from the middle of the capillary with one hand while holding the pipette in the other hand. The now exposed end of the capillary that was previously attached to the pipette tip and now facing upwards was then gently capped with another pre-cut 4 mm silicone cap. Finally, both silicone caps on either end of the capillary were gently squeezed towards each other to ensure a tight seal.

We rapidly thermal cycled the ddPCR droplets by placing the capped capillaries in a steel wire holder and manually moving the capillaries between the two adjacent pre-heated water baths. For the IDH R132H assay, we performed 45 cycles of denaturation (95 °C water bath) and 1 second of annealing/extension (62 °C water bath. For the BRAF V600E assay, we performed 40 cycles of denaturation (95 °C water bath) and 5 seconds of annealing/extension (62 °C water bath). We used a timer to determine when to switch between water baths. After thermal cycling, the capillary was held vertically and the cap facing upwards carefully removed by holding the capillary with one hand and pulling the cap with the other. The now uncapped end of the capillary was then attached to a new tip on a 200 μL pipette set to 47 μL. The silicone cap still attached to the capillary was then carefully removed by using one hand to apply pressure against the capillary towards the pipette tip, ensuring the seal is secure, holding the capillary between the index and middle finger, and pushing the cap off the capillary using the thumb while the pipette is held with the other hand. The capillary contents were then slowly dispensed into a ddPCR 96-well plate well. This unsealed ddPCR 96-well plate containing the amplified droplets was placed directly in the QX200 Droplet Reader and the amplified droplets were then read and analyzed as previously described.

### ddPCR sensitivity experiments

To determine the sensitivities of the ddPCR assays, we serially diluted IDH1 R132H and BRAF V600E 50% mutant DNA percentage (heterozygous, clonal) reference genomic DNA standards (Horizon Discovery) with non-mutant genomic DNA (NA12878 and NA12877 [Coriell], respectively) to create mutant DNA fractions of 25%, 10%, 5%, 1%, 0.1%, and 0.01%. We then measured the mutant DNA fractions of the 50% reference DNA and each dilution using standard ddPCR and UR-ddPCR. We performed this experiment twice for the IDH1 R132H assay, once with 32X Aptamer HS Taq concentration and once with 25X concentration. We performed this experiment for the BRAF V600E assay with 25X Aptamer HS Taq concentration.

### Tissue-to-result ultra-rapid ddPCR laboratory experiments

The tissue-to-result UR-ddPCR laboratory experiments aimed to achieve the fastest possible measurement of mutant DNA percentage and to confirm that UR-ddPCR measures mutant DNA percentages comparable to those measured by standard ddPCR. We do not include in the tissue-to-result time the four UR-DNA extraction preparation steps and the four UR-ddPCR preparation steps described above, because these steps are completed prior to tissue samples when conducting intraoperative UR-ddPCR.

Once the previously described preparation steps for UR-DNA extraction and UR-ddPCR were completed, we began the timers and either 1 or 4 pre-aliquoted and thawed IDH1 R132H mutant tumor tissue samples were processed per the previously described UR-DNA extraction protocol. Once the DNA was extracted, the timers were paused while 10 μL of the UR-DNA extract from each sample was aliquoted to a separate PCR tube stored on ice for later use in the standard ddPCR experiment. We then resumed the timers and conducted the UR-ddPCR assay as previously described, stopping the timers once the results were accessible on the QX200 droplet reader. During the subsequent analysis of UR-ddPCR results, the positive droplet thresholds were set by the experimenter, but the experimenter remained blind to the reported mutant DNA percentages of each of these samples until after the follow-up standard ddPCR experiment was completed. This ensured there was no bias when the experimenter set the positive droplet thresholds for the standard ddPCR samples.

Throughout the tissue-to-result UR-ddPCR experiment, the total time was measured on one timer and the times for each individual step of the process were measured using other separate timers that were present at each station (DNA extraction, droplet generation, droplet loading, PCR thermal cycling, droplet dispensing, and droplet reading). The time difference between the total time and the summed step-by-step times was the considered “movement time” and was excluded from the reported “tissue-to-result total time” since it is 1) dependent on the laboratory space in which the protocol is conducted, 2) could be reduced to 30 seconds or less if all machines were present at a single location, and 3) was only 1 minute and 47 seconds on average within our unoptimized laboratory space.

Immediately following each of these tissue-to-result UR-ddPCR experiments, we conducted a standard ddPCR assay as previously described for the same samples, with an input of 0.5 μL of the UR-DNA extract that was set aside after UR-DNA extraction. This blinded standard ddPCR measurement on the same DNA extract provided an assessment of the accuracy of our tissue-to-result UR-ddPCR.

### Tissue-to-result UR-ddPCR in the operating room

All patients whose samples were profiled as part of our intraoperative studies were consented under a human subjects protocol approved by the New York University Grossman School of Medicine Institutional Review Board. The research protocol required that the UR-ddPCR results remain unknown to the surgical team until after completion of the surgery, since this was the first research implementation of this method.

Three mobile carts were assembled for the operating room: one for UR-DNA extraction, one for pre-PCR UR-ddPCR steps, and one for post-PCR UR-ddPCR steps. The layout and all components used in each of these carts are detailed in **Supplementary** Figure 3 and **Supplementary Table 3**. Although all the carts could fit in the operating room, in order to conserve space, the carts were placed in a side room near the operating room.

In the morning prior to each surgical case, we prepared the following:

1. UR-ddPCR reaction mix: In cases # 1-3, this was prepared in two separate parts, A and B, that were stored at room temperature and combined immediately prior to each reaction in the operating room in an initial attempt to maximize reagent stability over the course of a prolonged surgery. We refer to this as intraoperative protocol version 1. Part A (7 μL/sample) contained the UR ddPCR IDH1 R132H assay primers and probes and part B (14 μL/sample) contained HaeIII, Aptamer HS Taq DNA Polymerase, and the ddPCR supermix for probes (no dUTP). Once parts A and B were combined, the composition of the UR-ddPCR reaction mix is identical to that used in UR-ddPCR laboratory experiments described above with 32x HS Taq concentration (0.8 U/μL) except with a final volume of 21 μL/reaction, since only 1 μL of DNA is added in the operating room assays. However, after obtaining lower positive droplet counts for UR-ddPCR in the operating room compared to UR-ddPCR in the laboratory, we altered the method of preparing UR-ddPCR reaction mix for the remaining cases # 4-13, which we refer to as intraoperative protocol version 2. For those cases, the UR-ddPCR reaction mix was prepared in the same way and with the same composition as used in UR-ddPCR laboratory experiments described above with 25x HS Taq concentration (0.625 U/μL), except with a final volume of 21 μL/reaction. The HS Taq concentration was lowered to 25x instead of 32x, since we found in the laboratory that high concentrations of polymerase can cause droplet instability. Notably, this change did not affect assay performance (**Supplemental Figure 1D**). 21 μL of the reaction mix was then pre-aliquoted into one well per strip tube, with a separate strip tube for each sample and control that will be run for the surgical case. During the surgical case, the pre-prepared strip tubes were stored at 4°C in a mini refrigerator until use.
2. 2. We added 22 μL of 1X ddPCR Buffer Control for Probes to all the remaining strip tube wells that would not be used for a tissue sample, i.e., the remaining 7 wells of each strip tube that do not contain the UR-ddPCR reaction mix.
3. We activated buffer ME as described for laboratory UR-ddPCR.
4. To conserve space in the operating room, we used a TissueLyser LT instrument (Qiagen) for homogenization with the 12-tube adaptor (Qiagen), instead of the TissueLyser II instrument that we used in the laboratory. We set the instrument to a 50 Hz, 30 second program.
5. We added 200 μL of activated buffer ME and six 2.8 mm ceramic beads (Omni, cat # 19-646-3) to each of a number of 2 mL DNA LoBind tubes (Eppendorf) corresponding to the anticipated number of tissue and control samples for that case. These tubes were stored at room temperature, and we confirmed in a tissue-to-result UR-ddPCR experiment that activated buffer ME is stable at room temperature for at least 3 hours. We used ceramic instead of steel beads for UR-ddPCR in the operating room, because these were found to homogenize tissue more effectively in the TissueLyser LT instrument.
6. We pre-heated a mini dry bath (Fisher, cat # 14-955-218) with a 2 mL adaptor (Fisher, cat # 14-955-225) to 98 °C for use in the UR-DNA extraction heat step.
7. We prepared the two 1-liter water baths, one at 95 °C and one at 62 °C.

After these preparation steps were completed, we performed two types of negative control assays. The first negative control assay to exclude reagent contamination was performed by following the UR-DNA extraction process without any tissue input followed by UR-ddPCR. Across all cases, this yielded 0% average mutant DNA percentage for both UR-ddPCR and standard ddPCR (N=12, since 2 of 13 cases were conducted on the same day and shared a set of control experiments). Note, we assigned a 0% mutant DNA percentage to negative control samples with ≤ 3 total positive droplets (i.e., non-mutant plus mutant positive droplets), because this low number of positive droplets yields unreliable estimates of mutant DNA percentage. The second negative control assay we performed measured the assay’s false positive rate using 1 μL of 30 ng/μL non-mutant DNA (NA12877) added to the UR-ddPCR reaction mix. Across all cases, this yielded an average mutant DNA percentage of 0.06% and 0.04% (N=12) in UR-ddPCR and standard ddPCR, respectively.

For cases 1-3 in which the UR-ddPCR mix was pre-prepared in two parts, approximately 5 to 10 minutes before each tissue sample was obtained in the operating room, we combined 7 μL of the UR-ddPCR mix part A and 14 μL of UR-ddPCR mix part B. For all cases, prior to each tissue sample becoming available, we also added 70 μL of Droplet Generation Oil for Probes to all DG8 cartridge oil wells, we initialized the QX Manager Software with the plate name and probes, and we configured wells in the software for analysis.

Upon resection of each approximately 8 x 8 x 2 mm tissue sample that we planned to profile in the operating room, the tissue was cut by the surgical team into two halves. One half was designated for profiling by the NIO Stimulated Raman Histology System^15^ (Invenio Imaging) followed by clinical pathology evaluation, and the other half was designated for UR-ddPCR. Additional tumor samples that were not profiled by ddPCR or NIO were also submitted for neuropathology evaluation.

Next, we performed UR-DNA extraction for each tissue sample by adding the tissue sample to the tube containing activated buffer ME and beads. UR-DNA extraction was then conducted per the UR-DNA extraction laboratory protocol except that homogenization used a TissueLyser LT instrument at 50 Hz for 30 seconds and the post-homogenization heat incubation was performed in the mini dry bath described above.

After UR-DNA extraction, we used 0.5 μL (case 1) or 1 μL (cases 2-13) of the UR-DNA extract as input into the UR-ddPCR reaction. The volume of UR-DNA extract was increased following case 1 to increase the numbers of positive droplets (mutant and non-mutant). We then followed the UR-ddPCR protocol as described above. For cases # 1-3, we used capillaries from Component Supply Company and for the remaining cases we used capillaries from Ziggy’s Tubes and Wires, as detailed in the ‘Ultra-rapid ddPCR in the laboratory’ section. Additionally, during the droplet generation step, 20 μL of the UR-DNA extract of each sample was saved in a 1.5 mL DNA LoBind tube (Eppendorf) for later profiling by standard ddPCR. The above process was repeated for each tissue sample as it was resected.

Throughout the intraoperative UR-ddPCR process, the step-by-step times were measured using timers present at each station. The total time was calculated as the time between beginning the bead homogenization to the time when results were available after the sample was read on the droplet reader. Note, we also measured the time of walking from the operating room to the side room, but this was excluded from the reported “total tissue-to-result time” since it is dependent on our specific operating room layout, and this time could be eliminated by placing all the instruments in the operating room.

Once the surgical day was completed, a positive control assay was performed by adding 1 μL of 30 ng/uL reference genomic DNA containing a 50% mutant DNA percentage (either IDH1 R132H or BRAF V600E depending on the case’s target assay) to the UR-ddPCR reaction mix instead of UR-DNA extract and performing the UR-ddPCR intraoperative protocol beginning at droplet generation. Across all cases, this yielded average mutant DNA percentages of 46% and 49% (N=12) in UR-ddPCR and standard ddPCR, respectively.

Finally, we conducted standard ddPCR in the laboratory for all of the controls and tissue samples of the case. This standard ddPCR was conducted as previously described using 1 μL input of either control DNA or UR-DNA extract.

Due to large error in estimating tumor cell percentages when there are low positive droplet counts, tissue samples with < 100 total positive droplets (i.e., IDH1 R132H-mutant plus IDH1 non-mutant positive droplets) were excluded from analysis and plots, though we list their results in **Supplementary Table 4**. This low positive droplet count can occur due to variability in tissue sample size and cellularity, and it occurred for only 1 of 50 samples in this study. For all intraoperative samples, we estimated tumor cell percentage as 2 x mutant DNA percentage. Specifically, the mutant DNA percentage is obtained from the “fractional abundance” field in the QuantaSoft software’s “data table” tab. The 95% Poisson confidence intervals for mutant DNA percentage were obtained from this data table’s “PoissonFractionalAbundanceMin” and “PoissonFractionalAbundanceMax” fields. The confidence interval for tumor cell percentage was then calculated as 2 x PoissonFractionalAbundanceMin to 2 X PossionFractionAbundanceMax. These tumor cell percentage estimates assume that, in all tumor cells, the assayed mutation is heterozygous and that the mutation locus has a copy number of 2, which is true for the large majority of IDH1 mutant tumors^5,32^ and for the majority of BRAF V600E mutant tumors^33^. Heterozygosity and absence of copy number changes was confirmed for all our IDH1 R132H mutant cases by subsequent clinical sequencing and methylation profiling (**Supplementary Table 4**). Future UR-ddPCR assays could feasibly also measure copy number of target loci^34^.

Additional samples from each case underwent neuropathology profiling by: 1) hematoxylin and eosin slides, 2) immunohistochemistry for IDH1 R132H (Dianova, GDIA-H09), 3) clinically-validated whole-genome DNA methylation profiling and classification with the Heidelberg Classifier v12 as previously described^35^, and 4) mutation and copy number analysis using the FDA-cleared NYU Langone Genome PACT matched tumor-normal DNA next-generation sequencing assay (FDA 510(k): K202304).

## Author Contributions

Z.R.M., D.A.O., and G.D.E. conceived of the project. Z.R.M. and G.D.E. designed the method development experiments. Z.R.M. performed the method development experiments. Z.R.M. and E.C.B. performed all other laboratory experiments. T.R. assisted with research subject coordination. Z.R.M., E.C.B., D.A.O., and G.D.E. designed the intraoperative experiments. E.B. performed intraoperative experiments with assistance from A.S, L.I.K., and D.R. Y.W. and M.S. contributed clinical and pathology data analyses. Z.R.M., E.C.B., D.A.O., and G.D.E. wrote the manuscript and all co-authors contributed to the final manuscript.

## Data Availability

All data produced in the present work are contained in the manuscript.

## Acknowledgements

This work was supported by discretionary laboratory start-up funds (G.D.E.) and by National Institutes of Health grant R01-CA226527 (D.A.O.). A QX200 system, reagents, and consumables were provided by Bio-Rad for the intraoperative phase of the project. Bio-Rad provided these items in-kind, without any input into the scientific conduct, analysis, or publication of this project.

## References

1 Samsom, K. G. et al. Feasibility of whole-genome sequencing-based tumor diagnostics in routine pathology practice. The Journal of Pathology 258, 179–188 (2022).

2 Ossowski, S. et al. Improving Time to Molecular Testing Results in Patients With Newly Diagnosed, Metastatic Non–Small-Cell Lung Cancer. JCO Oncology Practice 18, e1874–e1884 (2022).

3 Molinaro, A. M. et al. Association of Maximal Extent of Resection of Contrast-Enhanced and Non–Contrast-Enhanced Tumor With Survival Within Molecular Subgroups of Patients With Newly Diagnosed Glioblastoma. JAMA Oncology 6, 495–503 (2020).

4 Huang, L. E. Friend or foe—IDH1 mutations in glioma 10 years on. Carcinogenesis 40, 1299–1307 (2019).

5 Hartmann, C. et al. Type and frequency of IDH1 and IDH2 mutations are related to astrocytic and oligodendroglial differentiation and age: a study of 1,010 diffuse gliomas. Acta Neuropathologica 118, 469–474 (2009).

6 Di Nunno, V., Gatto, L., Tosoni, A., Bartolini, S. & Franceschi, E. Implications of BRAF V600E mutation in gliomas: Molecular considerations, prognostic value and treatment evolution. Frontiers in Oncology 12 (2023).

7 Dankner, M., Rose, A. A. N., Rajkumar, S., Siegel, P. M. & Watson, I. R. Classifying BRAF alterations in cancer: new rational therapeutic strategies for actionable mutations. Oncogene 37, 3183–3199 (2018).

8 Gerstung, M. et al. The evolutionary history of 2,658 cancers. Nature 578, 122–128 (2020).

9 Suzuki, H. et al. Mutational landscape and clonal architecture in grade II and III gliomas. Nature Genetics 47, 458–468 (2015).

10 Delev, D. et al. Surgical management of lower-grade glioma in the spotlight of the 2016 WHO classification system. Journal of Neuro-Oncology 141, 223–233 (2019).

11 Santamaria-Barria, J. A. & Mammen, J. M. V. Surgical Management of Melanoma: Advances and Updates. Current Oncology Reports 24, 1425–1432 (2022).

12 Sanai, N. & Berger, M. S. Surgical oncology for gliomas: the state of the art. Nature Reviews Clinical Oncology 15, 112–125 (2018).

13 Orringer, D. et al. Extent of resection in patients with glioblastoma: limiting factors, perception of resectability, and effect on survival: Clinical article. Journal of Neurosurgery JNS 117, 851–859 (2012).

14 Beiko, J. et al. IDH1 mutant malignant astrocytomas are more amenable to surgical resection and have a survival benefit associated with maximal surgical resection. Neuro Oncol 16, 81–91 (2013).

15 Hollon, T. C. et al. Near real-time intraoperative brain tumor diagnosis using stimulated Raman histology and deep neural networks. Nature Medicine 26, 52–58 (2020).

16 Shankar, G. M. et al. Rapid Intraoperative Molecular Characterization of Glioma. JAMA Oncology 1, 662–667 (2015).

17 Wadden, J. et al. Ultra-rapid somatic variant detection via real-time targeted amplicon sequencing. Communications Biology 5, 708 (2022).

18 Farrar, J. S. & Wittwer, C. T. Extreme PCR: efficient and specific DNA amplification in 15–60 seconds. Clinical chemistry 61, 145–153 (2015).

19 Chang, M. T. et al. Identifying recurrent mutations in cancer reveals widespread lineage diversity and mutational specificity. Nature Biotechnology 34, 155–163 (2016).

20 Vermeulen, C. et al. Ultra-fast deep-learned CNS tumour classification during surgery. Nature 622, 842–849 (2023).

21 Feng, Z. et al. Rapid detection of isocitrate dehydrogenase 1 mutation status in glioma based on Crispr-Cas12a. Scientific Reports 13, 5748 (2023).

22 Wainman, L. M., Sathyanarayana, S. H. & Lefferts, J. A. Applications of Digital Polymerase Chain Reaction (dPCR) in Molecular and Clinical Testing. The Journal of Applied Laboratory Medicine 9, 124–137 (2024).

23 Youssef, G. & Miller, J. J. Lower Grade Gliomas. Current Neurology and Neuroscience Reports 20, 21 (2020).

24 Satgunaseelan, L. et al. Prognostic and predictive biomarkers in central nervous system tumours: the molecular state of play. Pathology 56, 158–169 (2024).

25 Hadjipanayis, C. G. & Stummer, W. 5-ALA and FDA approval for glioma surgery. Journal of neuro-oncology 141, 479–486 (2019).

26 van Dam, G. M. et al. Intraoperative tumor-specific fluorescence imaging in ovarian cancer by folate receptor-α targeting: first in-human results. Nature Medicine 17, 1315–1319 (2011).

27 Tanyi, J. L. et al. A Phase III Study of Pafolacianine Injection (OTL38) for Intraoperative Imaging of Folate Receptor–Positive Ovarian Cancer (Study 006). Journal of Clinical Oncology 41, 276–284 (2022).

28 Nasrallah, M. P. et al. Machine learning for cryosection pathology predicts the 2021 WHO classification of glioma. Med 4, 526–540.e524 (2023).

29 Shen, J. et al. A rapid nucleic acid concentration measurement system with large field of view for a droplet digital PCR microfluidic chip. Lab on a Chip 21, 3742–3747 (2021).

30 Ren, Y. et al. A three-in-one microfluidic droplet digital PCR platform for absolute quantitative analysis of DNA. Lab on a Chip (2023).

31 Untergasser, A. et al. Primer3—new capabilities and interfaces. Nucleic Acids Research 40, e115–e115 (2012).

32 The-Cancer-Genome-Atlas-Research-Network. Comprehensive, Integrative Genomic Analysis of Diffuse Lower-Grade Gliomas. New England Journal of Medicine 372, 2481–2498 (2015).

33 Hélias-Rodzewicz, Z. et al. Variations of BRAF mutant allele percentage in melanomas. BMC Cancer 15, 497 (2015).

34 Wolter, M., Felsberg, J., Malzkorn, B., Kaulich, K. & Reifenberger, G. Droplet digital PCR-based analyses for robust, rapid, and sensitive molecular diagnostics of gliomas. Acta Neuropathologica Communications 10, 42 (2022).

35 Galbraith, K. et al. Clinical utility of whole-genome DNA methylation profiling as a primary molecular diagnostic assay for central nervous system tumors— A prospective study and guidelines for clinical testing. Neuro-Oncology Advances 5 (2023).

