## Supplementary figures and images for "Ultra-Rapid Droplet Digital PCR Enables Intraoperative Tumor Quantification"

### Supplementary Figure 1

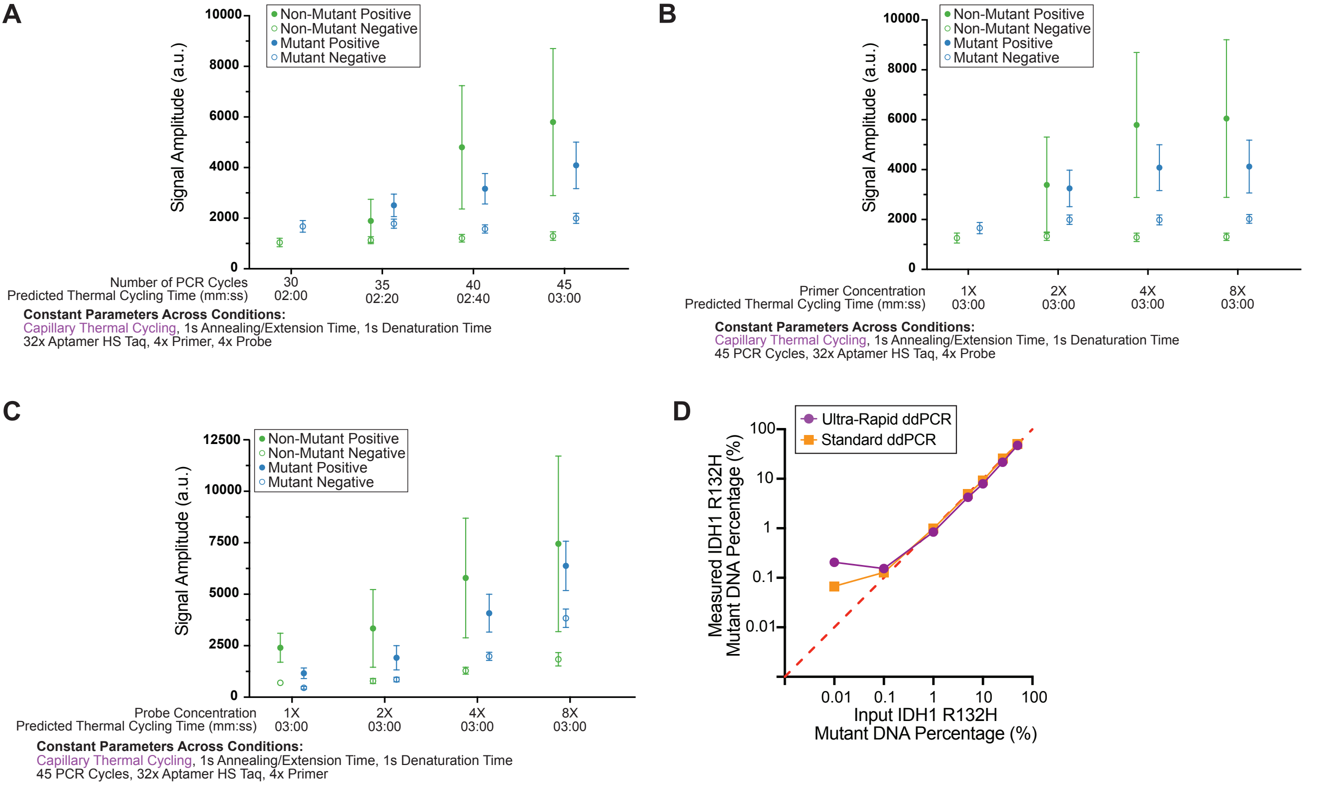

### Supplementary Figure 2

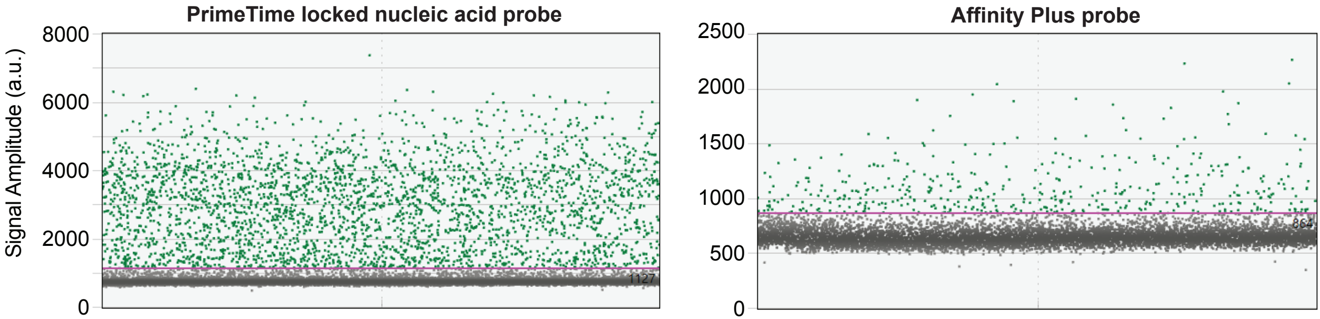

### Supplementary Figure 3

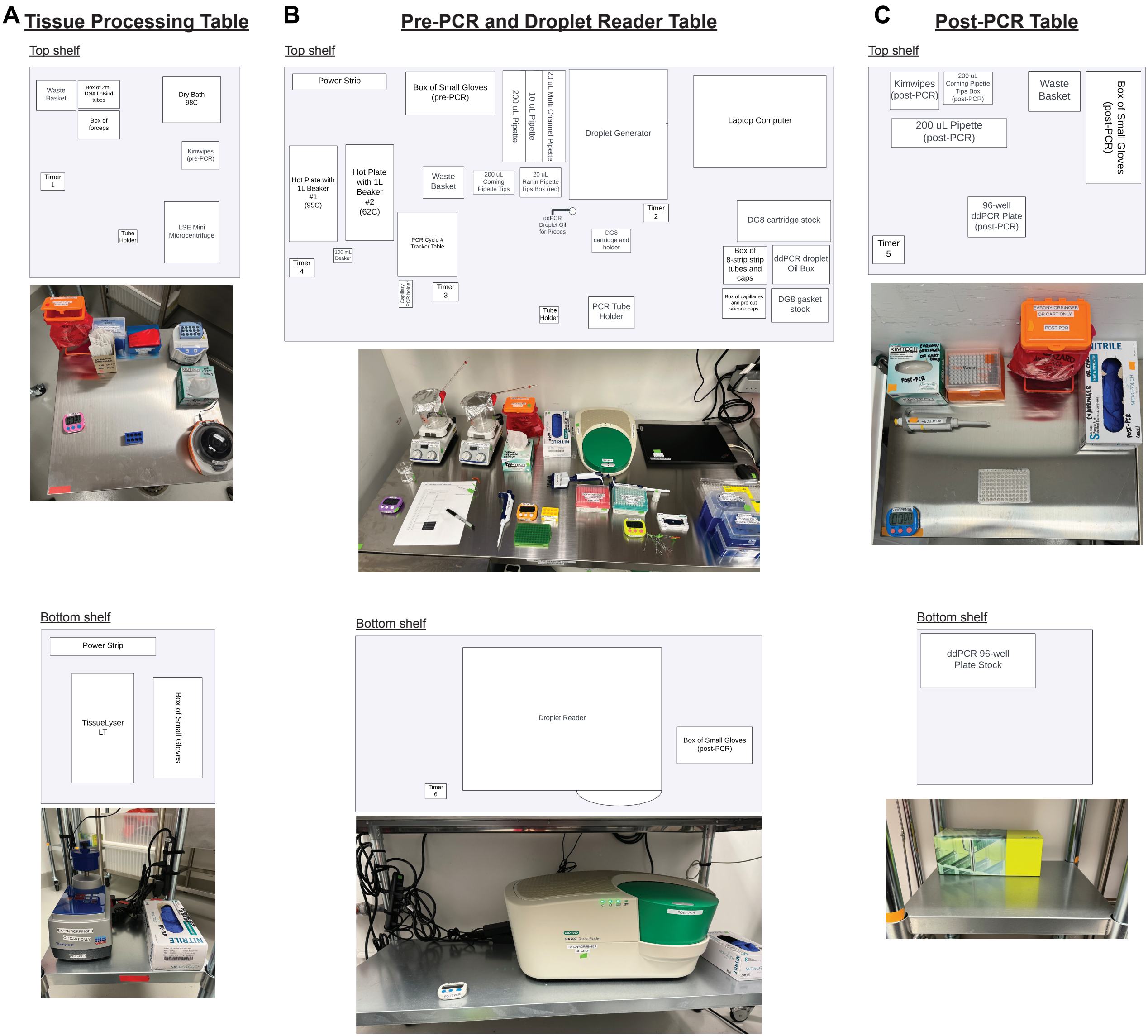

### Supplementary Figure 4

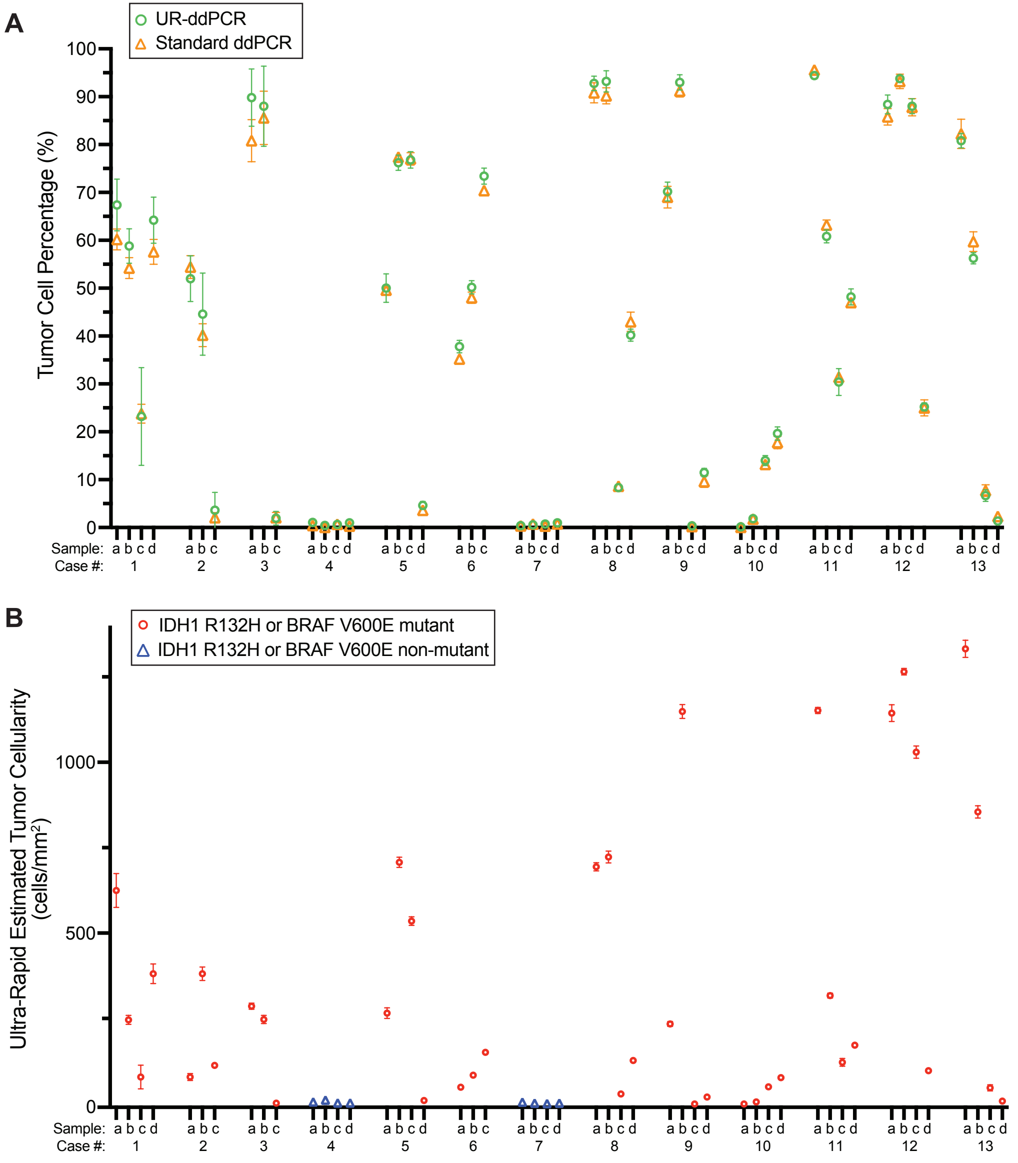
