## Supplementary File for "Ultra-Rapid Droplet Digital PCR Enables Intraoperative Tumor Quantification"

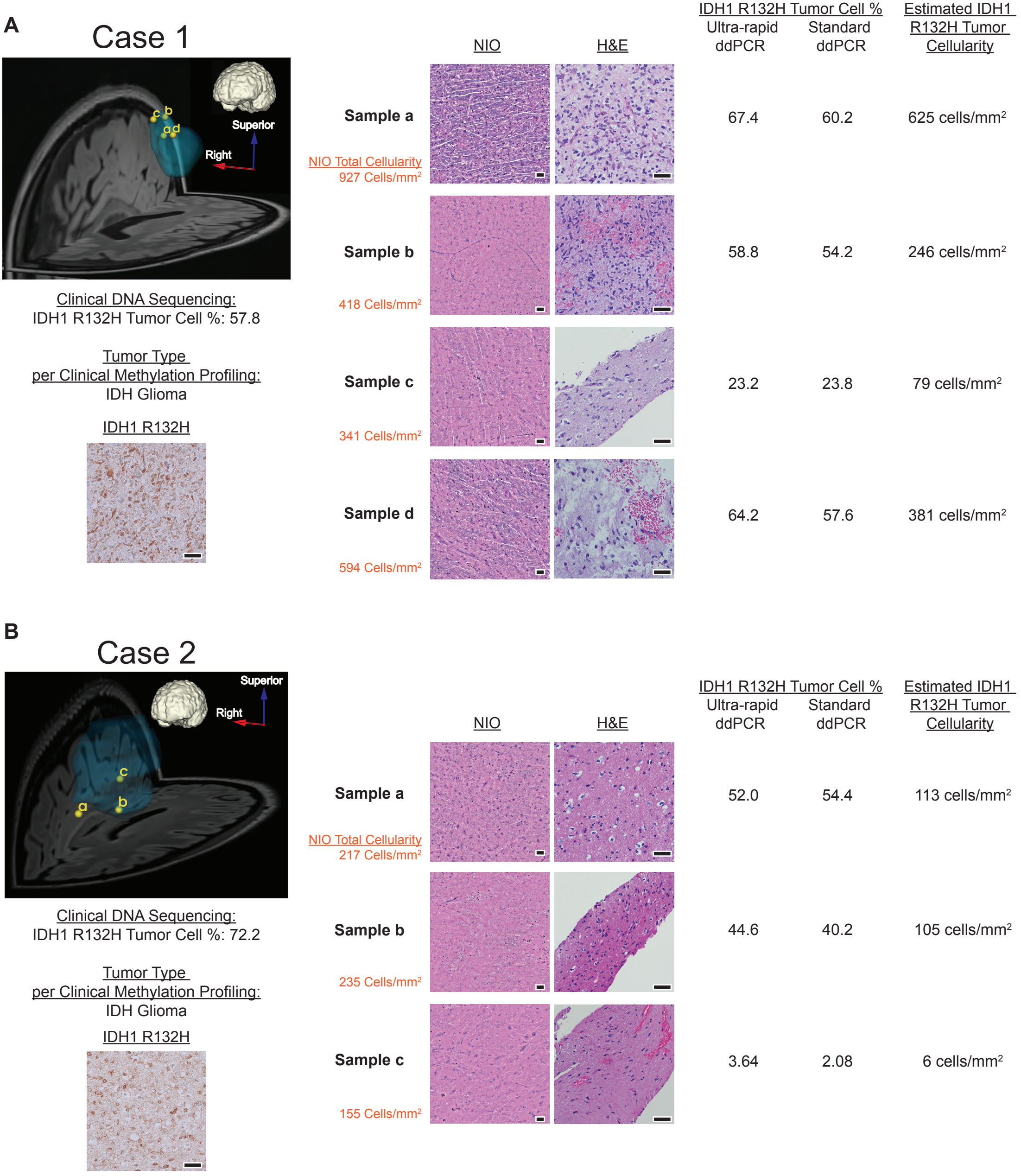
A
Case 1
Superior
Right
c b
a d
Clinical DNA Sequencing:
IDH1 R132H Tumor Cell %: 57.8
Tumor Type
per Clinical Methylation Profiling:
IDH Glioma
IDH1 R132H
NIO
H&E
IDH1 R132H Tumor Cell %
Ultra-rapid ddPCR
Standard ddPCR
Estimated IDH1 R132H Tumor Cellularity
Sample a
NIO Total Cellularity
927 Cells/mm²
67.4
60.2
625 cells/mm²
Sample b
418 Cells/mm²
58.8
54.2
246 cells/mm²
Sample c
341 Cells/mm²
23.2
23.8
79 cells/mm²
Sample d
594 Cells/mm²
64.2
57.6
381 cells/mm²
B
Case 2
Superior
Right
c
a
b
Clinical DNA Sequencing:
IDH1 R132H Tumor Cell %: 72.2
Tumor Type
per Clinical Methylation Profiling:
IDH Glioma
IDH1 R132H
NIO
H&E
IDH1 R132H Tumor Cell %
Ultra-rapid ddPCR
Standard ddPCR
Estimated IDH1 R132H Tumor Cellularity
Sample a
NIO Total Cellularity
217 Cells/mm²
52.0
54.4
113 cells/mm²
Sample b
235 Cells/mm²
44.6
40.2
105 cells/mm²
Sample c
155 Cells/mm²
3.64
2.08
6 cells/mm²

**C**

### Case 3

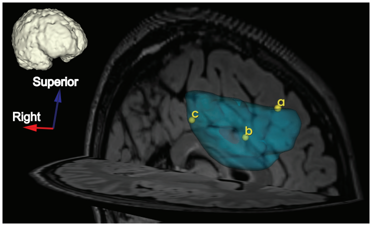


Clinical DNA Sequencing:
IDH1 R132H Tumor Cell %: 77.6

Tumor Type
per Clinical Methylation Profiling:
IDH Glioma

IDH1 R132H

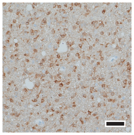

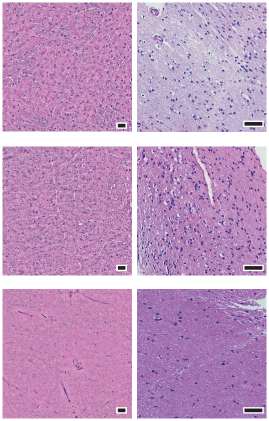


| | NIO Total Cellularity | IDH1 R132H Tumor Cell % Ultra-rapid ddPCR | IDH1 R132H Tumor Cell % Standard ddPCR | Estimated IDH1 R132H Tumor Cellularity |
|---|---|---|---|---|
| Sample a | 319 Cells/mm² | 89.8 | 80.8 | 286 cells/mm² |
| Sample b | 281 Cells/mm² | 88.0 | 85.6 | 247 cells/mm² |
| Sample c | 104 Cells/mm² | 1.96 | 2.06 | 2 cells/mm² |

**D**

### Case 4

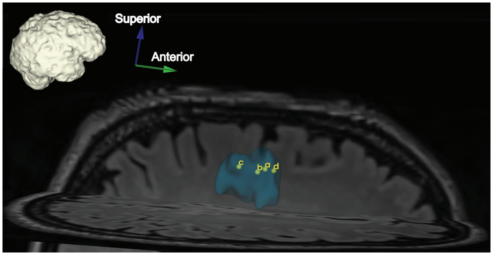


Clinical DNA Sequencing:
IDH1 R132H Tumor Cell %: 0

Tumor Type
per Clinical Methylation Profiling:
IDH Wildtype Glioblastoma

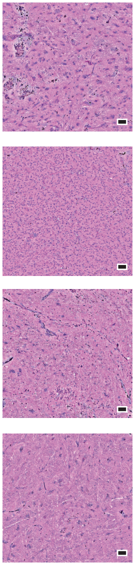


| | NIO Total Cellularity | IDH1 R132H Tumor Cell % Ultra-rapid ddPCR | IDH1 R132H Tumor Cell % Standard ddPCR | Estimated IDH1 R132H Tumor Cellularity |
|---|---|---|---|---|
| Sample a | 529 Cells/mm² | 1.06 | 0.42 | 6 cells/mm² |
| Sample b | 2585 Cells/mm² | 0.43 | 0.06 | 11 cells/mm² |
| Sample c | 477 Cells/mm² | 0.63 | 0.71 | 3 cells/mm² |
| Sample c | 272 Cells/mm² | 0.98 | 0.33 | 3 cells/mm² |

**E**

### Case 5

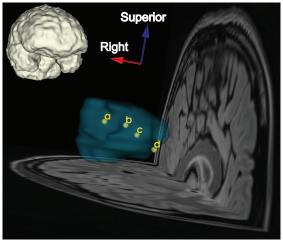


Clinical DNA Sequencing:
IDH1 R132H Tumor Cell %: 58.6

Tumor Type per Clinical Methylation Profiling:
IDH Glioma

IDH1 R132H

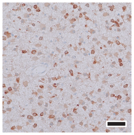

NIO | H&E

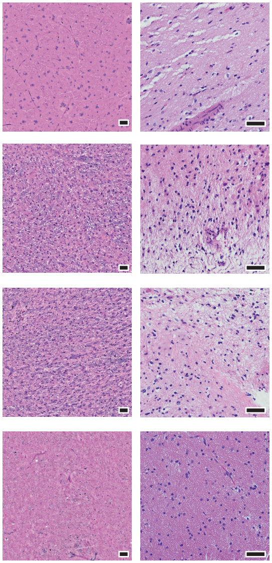

| Sample | NIO Total Cellularity | IDH1 R132H Tumor Cell % Ultra-rapid ddPCR | IDH1 R132H Tumor Cell % Standard ddPCR | Estimated IDH1 R132H Tumor Cellularity |
|---|---|---|---|---|
| Sample a | 532 Cells/mm$^2$ | 50.0 | 49.6 | 266 cells/mm$^2$ |
| Sample b | 928 Cells/mm$^2$ | 76.2 | 77.4 | 707 cells/mm$^2$ |
| Sample c | 696 Cells/mm$^2$ | 76.8 | 77.0 | 535 cells/mm$^2$ |
| Sample d | 225 Cells/mm$^2$ | 4.62 | 3.62 | 10 cells/mm$^2$ |

**F**

### Case 6

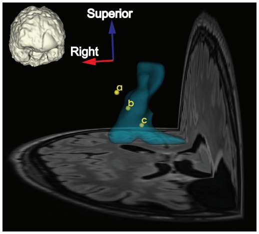


Clinical DNA Sequencing:
IDH1 R132H Tumor Cell %: 64.6

Tumor Type per Clinical Methylation Profiling:
IDH Glioma

NIO | H&E

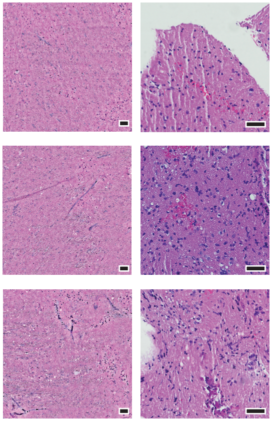

| Sample | NIO Total Cellularity | IDH1 R132H Tumor Cell % Ultra-rapid ddPCR | IDH1 R132H Tumor Cell % Standard ddPCR | Estimated IDH1 R132H Tumor Cellularity |
|---|---|---|---|---|
| Sample a | 129 Cells/mm$^2$ | 37.8 | 35.2 | 49 cells/mm$^2$ |
| Sample b | 167 Cells/mm$^2$ | 50.2 | 48.0 | 84 cells/mm$^2$ |
| Sample c | 206 Cells/mm$^2$ | 73.4 | 70.4 | 151 cells/mm$^2$ |

**G**

### Case 7

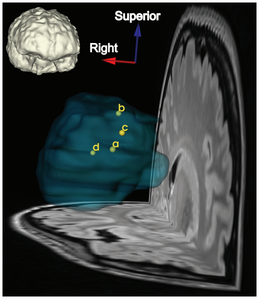


Clinical DNA Sequencing:
IDH2 R172M %: 85.9

Tumor Type
per Clinical Methylation Profiling:
IDH Glioma

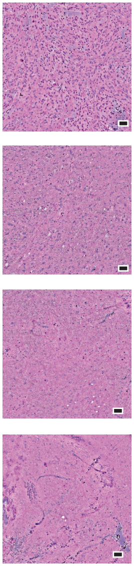

| | NIO Total Cellularity | NIO | IDH1 R132H Tumor Cell % Ultra-rapid ddPCR | IDH1 R132H Tumor Cell % Standard ddPCR | Estimated IDH1 R132H Tumor Cellularity |
|---|---|---|---|---|---|
| **Sample a** | 1439 Cells/mm$^2$ | | 0.38 | 0.35 | 5 cells/mm$^2$ |
| **Sample b** | 295 Cells/mm$^2$ | | 0.58 | 0.74 | 2 cells/mm$^2$ |
| **Sample c** | 203 Cells/mm$^2$ | | 0.73 | 0.33 | 1 cells/mm$^2$ |
| **Sample d** | 170 Cells/mm$^2$ | | 0.97 | 0.79 | 2 cells/mm$^2$ |

**H**

### Case 8

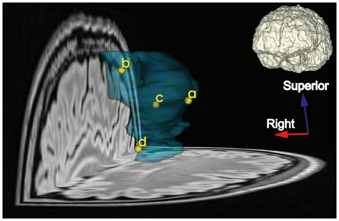


Clinical DNA Sequencing:
IDH1 R132H Tumor Cell %: 89.0

Tumor Type
per Clinical Methylation Profiling:
IDH Glioma

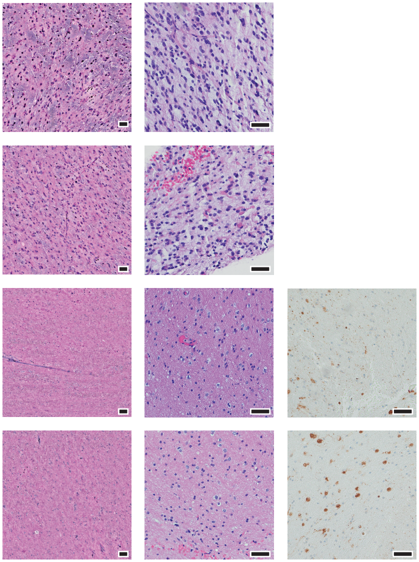

| | NIO Total Cellularity | NIO | H&E | IDH1 R132H | IDH1 R132H Tumor Cell % Ultra-rapid ddPCR | IDH1 R132H Tumor Cell % Standard ddPCR | Estimated IDH1 R132H Tumor Cellularity |
|---|---|---|---|---|---|---|---|
| **Sample a** | 748 Cells/mm$^2$ | | | | 92.8 | 90.8 | 694 cells/mm$^2$ |
| **Sample b** | 776 Cells/mm$^2$ | | | | 93.2 | 90.2 | 723 cells/mm$^2$ |
| **Sample c** | 351 Cells/mm$^2$ | | | | 8.36 | 8.66 | 29 cells/mm$^2$ |
| **Sample d** | 316 Cells/mm$^2$ | | | | 40.2 | 43.0 | 127 cells/mm$^2$ |

**I**

### Case 9

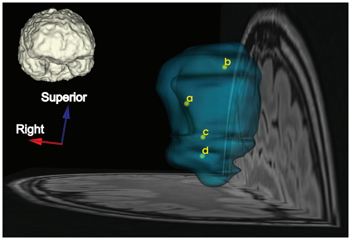


Clinical DNA Sequencing:
IDH1 R132H %: 67.6

Tumor Type
per Clinical Methylation Profiling:
IDH Glioma

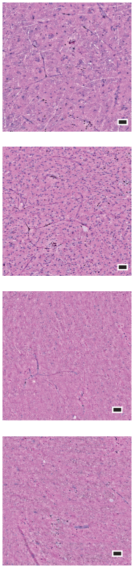

| | NIO Total Cellularity | IDH1 R132H Tumor Cell % Ultra-rapid ddPCR | IDH1 R132H Tumor Cell % Standard ddPCR | Estimated IDH1 R132H Tumor Cellularity |
|---|---|---|---|---|
| Sample a | 334 Cells/mm$^2$ | 70.2 | 69.0 | 234 cells/mm$^2$ |
| Sample b | 1236 Cells/mm$^2$ | 93.0 | 91.2 | 1149 cells/mm$^2$ |
| Sample c | 95 Cells/mm$^2$ | 0.37 | 0.24 | 0 cells/mm$^2$ |
| Sample d | 172 Cells/mm$^2$ | 11.5 | 9.60 | 20 cells/mm$^2$ |

**J**

### Case 10

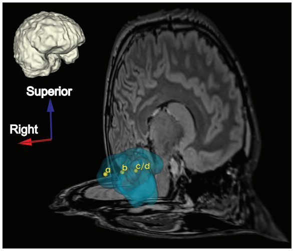


Clinical DNA Sequencing:
IDH1 R132H Tumor Cell %: 16.1,
matches sample d

Tumor Type
per Clinical Methylation Profiling:
IDH Astrocytoma

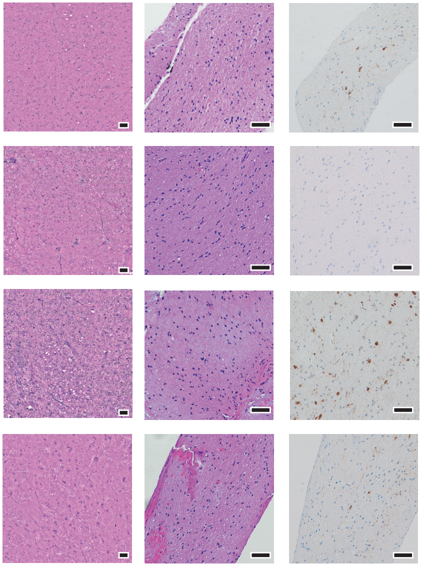


| | NIO Total Cellularity | IDH1 R132H Tumor Cell % Ultra-rapid ddPCR | IDH1 R132H Tumor Cell % Standard ddPCR | Estimated IDH1 R132H Tumor Cellularity |
|---|---|---|---|---|
| Sample a | 356 Cells/mm$^2$ | 0.14 | 0.08 | 0 cells/mm$^2$ |
| Sample b | 307 Cells/mm$^2$ | 1.85 | 1.79 | 6 cells/mm$^2$ |
| Sample c | 354 Cells/mm$^2$ | 14.0 | 13.2 | 50 cells/mm$^2$ |
| Sample d | 390 Cells/mm$^2$ | 19.7 | 17.8 | 77 cells/mm$^2$ |

**K**

### Case 12

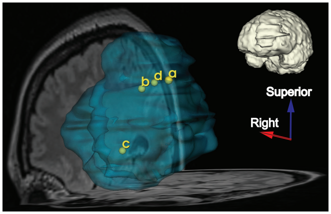


Clinical DNA Sequencing:
IDH1 R132H Tumor Cell %: 79.2

Tumor Type
per Clinical Methylation Profiling:
IDH Glioma

IDH1 R132H

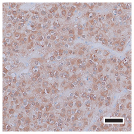

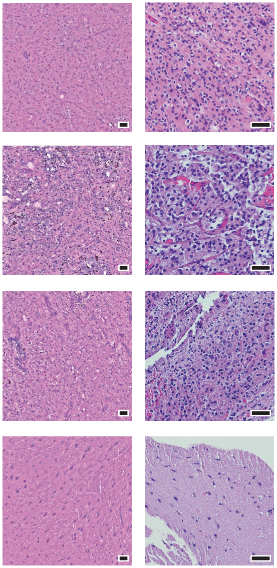

| Sample | NIO Total Cellularity | IDH1 R132H Tumor Cell % Ultra-rapid ddPCR | IDH1 R132H Tumor Cell % Standard ddPCR | Estimated IDH1 R132H Tumor Cellularity |
|---|---|---|---|---|
| Sample a | 1294 Cells/mm$^2$ | 88.4 | 85.8 | 1144 cells/mm$^2$ |
| Sample b | 1349 Cells/mm$^2$ | 93.8 | 93.2 | 1265 cells/mm$^2$ |
| Sample c | 1170 Cells/mm$^2$ | 88.0 | 87.8 | 1030 cells/mm$^2$ |
| Sample d | 386 Cells/mm$^2$ | 25.2 | 25.0 | 97 cells/mm$^2$ |

**L**

### Case 13

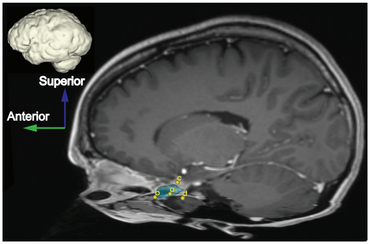


Clinical DNA Sequencing:
Data pending pathology

Tumor Type
per Clinical Methylation Profiling:
Data pending pathology

Immunohistochemistry of Prior Resection:
Plemorphic xanthoastrocytoma
BRAF V600E positive

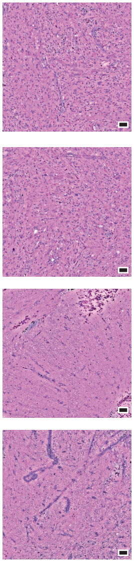

| Sample | NIO Total Cellularity | BRAF V600E Tumor Cell % Ultra-rapid ddPCR | BRAF V600E Tumor Cell % Standard ddPCR | Estimated BRAF V600E Tumor Cellularity |
|---|---|---|---|---|
| Sample a | 1648 Cells/mm$^2$ | 80.8 | 82.2 | 1332 cells/mm$^2$ |
| Sample b | 1519 Cells/mm$^2$ | 56.3 | 59.7 | 855 cells/mm$^2$ |
| Sample c | 708 Cells/mm$^2$ | 6.66 | 7.72 | 47 cells/mm$^2$ |
| Sample d | 563 Cells/mm$^2$ | 1.38 | 2.32 | 8 cells/mm$^2$ |
