## Supplementary Note for "Ultra-Rapid Droplet Digital PCR Enables Intraoperative Tumor Quantification"

**Supplementary Note: Comparison of standard and ultra-rapid ddPCR thermal conductance and surface area to volume ratios**

Here, we estimate the thermal conductances of the ddPCR 96-well plate well used in standard ddPCR and the stainless-steel capillary used in ultra-rapid ddPCR. Thermal conductance, C, is the rate at which a material of a given thickness conducts heat. It is calculated as C = K/T and measured in Watts/(meter$^2$*Kelvin), i.e., W/(m$^2$*K), where K is the thermal conductivity (a heat transfer constant specific to a given material) measured in Watts/(meter*Kelvin) and T is the thickness of the material, measured in meters.

The thermal conductivity of polypropylene, the plastic used in the ddPCR 96-well plates, has been measured to be 0.22 W/(m*K) [1], and the thermal conductivity of stainless-steel 304, the material of our capillaries, is approximately 15 W/(m*K) [2.] Since the thickness of the ddPCR plate wells was not available to us, we measured the thickness of 33 wells of a ddPCR 96-well plate at their thinnest point using a micrometer and obtained an estimate of 0.30 $\pm$ 0.07 mm. The thickness of the stainless-steel capillary is 0.10 $\pm$ 0.04 mm per the manufacturer. Thus, the thermal conductance of the ddPCR plate well is approximately 7.3 $\times 10^2$ W/(m$^2$*K) and the thermal conductance for the stainless-steel capillary is approximately 1.5 $\times 10^5$ W/(m$^2$*K), which is an ~ 204-fold difference.

We also estimated the surface area to volume ratio of a standard ddPCR reaction occurring in a ddPCR 96-well plate and an ultra-rapid ddPCR reaction occurring in a stainless steel capillary. We estimated the volume of the slanted portion of an individual well of a ddPCR 96-well plate as a truncated cone using bottom and top diameters of 2.16 mm and 5.60 mm, respectively, and a height of 11.10 mm as estimated from the manufacturer's blueprint, which yielded a volume of 139.84 µL. Using these values, we also calculated the surface area of the slanted portion of the ddPCR plate well to be 136.92 mm$^2$. Thus, the surface area to volume ratio of a standard ddPCR well is 0.98 mm$^2$/µL. For the stainless-steel capillary, we calculated its volume as a cylinder with length 57.15 mm and an inner diameter of 1.067 mm (per the manufacturer), which yielded 51.10 µL. Using these values, we also calculated the outer surface area of the capillary, which is the area contacting the heated water during ultra-rapid ddPCR, to be 191.57 mm$^2$. Thus, the surface area to volume ratio of the capillary is 3.75 mm$^2$/µL, which is 3.8-fold higher than that of the ddPCR plate well.

References


1 Antonella, P. & Domenico, A. in *Polypropylene* (eds Wang Weiyu & Zeng Yiming) Ch. 3 (IntechOpen, 2019).

2 Graves, R. S., Kollie, T. G., McElroy, D. L. & Gilchrist, K. E. The thermal conductivity of AISI 304L stainless steel. *International Journal of Thermophysics* **12**, 409-415 (1991).
